# Verification of the semiquantitative assessment of vertebral deformity for subsequent osteoporotic fracture prediction and screening for the initiation of osteoporosis treatment: A case-control study using a clinical-based setting

**DOI:** 10.1101/2024.12.22.24319509

**Authors:** Ichiro Yoshii, Naoya Sawada, Tatsumi Chijiwa

**Affiliations:** Department of Rheumatology and Musculoskeletal Medicine, Yoshii Clinic Postal address: 6-7-5 Nakamura-Ohashidori, Shimanto-City, 787-0033 Kochi Prefecture, Japan; Department of Rheumatology, Dohgo Onsen Hospital, Matsuyama, Ehime Prefecture, Japan; Department of Rheumatology, Kochi Memorial Hospital, Kochi, Kochi Prefecture, Japan

**Author notes:** **Authors’ contribution:** Ichiro Yoshii: Conceptualization, Part of Methodology, Software, Validation, Formal analysis, Investigation, Resources, Data Curation, Writing – Original Draft, Visualization, Project administration. Naoya Sawada: Data Curation, Validation, Recorded data, Writing - review & editing. Tatsumi Chijiwa: Data Curation, Validation, Recorded data, Writing - review & editing. **Ethical statement** The study protocols and patient consent requirements were approved by the Ethics Committee of the Genyu Medical Corporation (approval number: G-2023-3). The subjects and their families were informed that the personal information obtained in this study was anonymous and would only be used for analysis. **Consent for publication:** None. **Availability of data and material:** The datasets used and analyzed during the current study are available from the corresponding author upon reasonable request. **Code availability:** Not applicable. **English editing support and Use of AI application statement** The authors ordered a grammar check for the professional English editor who works with Enago, an English editing company. They did not use AIGC tools to assist with writing.

**Keywords:** incident fracture, prevalent fracture, semiquantitative grade, vertebral deformity, osteoporotic fracture, propensity score matching

## Abstract

**Background:** Semiquantitative grading of the vertebral body (SQ) is an easy screening method for vertebral body deformation. The validity of the SQ as a risk factor and as a screening tool for incident osteoporotic fractures (OF) was investigated using retrospective case-control data

**Methods:** Outpatients followed up for ≥ two years as patients with osteoporosis were recruited. All of them were tested with X-ray images of the lateral thoracolumbar view and other tests at baseline. Patients were classified according to the SQ grade, and potential risk factors were compared for each SQ group. Cox regression analyses were conducted regarding the incident OFs separated into vertebral and non-vertebral fractures (VF and NVF). Statistical differences in the possible risk factors among the groups were examined, and the likelihood of the incident OFs in the variables was examined. After propensity score matching procedures of the confounding factors (PSM), the possibility of incident VFs was compared between the SQ grade groups.

**Results:** In the crude dataset, the probabilities of incident VF in SQ Grade 3 and NVF in SQ Grade 1 or higher were significantly higher than in the other grades. Using a Cox regression analysis multivariant mode, the SQ grade was the only statistically significant factor for an incident VF, but not for NVF. However, no significant difference was shown after PSM for VFs.

**Conclusions:** These results suggested that the SQ classification was inappropriate for predicting an incident OFs. However, the grading showed significantly higher risk, therefore that is available for screening.

## Introduction

Osteoporosis is a disease that forms an internal bone environment prone to fracture due to weakening bone strength [1,2]. Risk factors for bone fragility include decreased bone mineral density [3], prolonged, chronic, and severe lifestyle-related diseases [4,5], low striated muscle mass [6], accessible falls caused by various locomotive diseases [7–11], decreased renal function [12], and poor nutritional\ status [10,13,14]. Typical osteoporotic fracture sites include the vertebral body, proximal femur, distal radius, and proximal humerus, of which vertebral fracture is among the most common [9,15,16]. Many cases of vertebral fracture (VF) result in fractures without even knowing the mechanism of injury. It is also known as a fracture, often found in the so-called Japanese “fracture before you know it,” in which the fracture occurs without an episodic fall [17]. Vertebral body fracture is a severe consequence of osteoporosis and is well-known primarily as one of the most serious risk factors for subsequent osteoporotic fracture [16,18,19]. Therefore, confirming the presence or history of VF through screening is very meaningful in preventing secondary vertebral fractures.

The severity of vertebral deformities and the risk of subsequent fractures are often discussed. In spine radiographs, the semiquantitative (SQ) criteria proposed by Genant et al. are commonly used to identify vertebral deformity (VD) in spines T4 to L4 [20]. Genant et al. classified the spine as usual by visual inspection of a lateral view of the spine X-ray image.

Because of its simplicity, the SQ method is often used to screen for VD. In Japan, some facilities have adopted it as a method of osteoporosis screening due to its visual easiness [21–23]. There is also a report that the evaluation of VD is a predictor of VF and is beneficial [24]. Yoshimura et al. conducted a large cohort study in three rural towns in Japan. They concluded that prevalent vertebral fracture is a risk factor for subsequent vertebral fractures, even in the Grade 1 minor deformity group [25]. Grade 2∼3 minor deformities are more common in men in the Genant SQ method [25,26]. However, these analyses did not describe the influence of confounding factors. Another report conducted a cross-sectional study regarding the relationship between mild wedge deformity and prevalence in vertebrae and found that mild vertebral deformities are likely to be the downtrend bone metabolism by aging. They may be deformities provoked not by osteoporosis but by aging-related spine diseases [27]. There are many confounding factors for assessing the prediction of subsequent VDs using the SQ method.

Little information exists on the association between SQ and osteoporotic non-vertebral fracture (NVF) [28]. However, if SQ is representative of a vertebral fracture, there should be a reasonable causal relationship between them.

We have investigated the frequency of incident osteoporotic fractures and risk weight of factors for the incident fractures using small retrospective case-control study data in a rural area in Japan.

## Materials and Methods

### Patient inclusion and exclusion

A lateral view of the thoracolumbar spine X-ray picture was taken of patients who consulted outpatients due to some musculoskeletal problem, and bone mineral density (BMD) was measured in the lumbar spine and femoral neck using dual-energy X-ray absorptiometry (DXA) with the DPX^®^ Bravo ME9309 Bone Densitometer (GE Health Care, Chicago, IL, USA: Coefficients of variation; CV: 1.1% (lumbar spine), 0.9% (femoral neck)) at the same time from March 2013 to December 2020 was picked up.

The date when the X-ray picture was taken was set as a baseline, and they were followed up at the shortest 24 months unless an incident fracture had developed. They also took blood samples for testing at the baseline. Patients who had failed to examine items used in the study were excluded. Patients who lost follow-up before two years due to some reasons without fractures, such as long-term admission because of severe comorbidities, move to another place, admission to a nursing home, and censored death, were excluded. Patients who had Scheuermann’s deformity and the presence of hypophosphatemic rickets were also extruded. In the included patients, follow-up periods were determined from the baseline to the last consult date. Patients who were censored incident vertebral fracture that was confirmed with or without an apparent injury episode but with any symptoms were examined by the lateral view of the spine X-ray picture; the date of the fracture was set as the last date. The SQ method was not used in these cases, but a change from the X-ray picture taken at the baseline was adopted. In the patients who suffered death, were admitted due to severe comorbidities, or were lost to follow-up due to unknown reasons after two years of follow-up, the last consult date to our institute was set as the latest.

### Primary vertebral deformity defining using the SQ method

Patients’ primary vertebral deformities were classified using the semiquantitative method (SQ) from the lateral view of the thoracolumbar spine X-ray picture. We arranged vertebral deformity following the identification of Genant’s SQ classification in vertebral height, that was, compression rate of the vertebral height from Grade 0 to Grade 3 (G-0, compression up to 5% in height; G-1, compression up to 25% or 10∼20% reduction in area; G-2, compression from 26 to 40% in height or 20–40% reduction in area; G-3, compression more than 40% in height and area). The two physicians of the authors performed the assessment. If the assessment did not match, the third assessor was assessed, and a majority vote made the decision. When multiple fractures were found in the X-ray picture, a more severe Grade was adopted. The Cohen Kappa score was determined.

### Other Factors extraction

Prevalent clinical VF and NVF that were extracted from the medical record, current smoke habitat, alcohol habitat, and parent’s fracture history were harvested from the interview at baseline. Positive findings with vertebral endplate collapse (VEC) using an algorithm-based quantitative method (ABQ), as observed with X-ray pictures, were defined from the lateral view of the thoracolumbar X-ray picture [30]. The presence of abdominal aortic calcification (AAC) was also determined from the lateral view of the thoracolumbar X-ray picture. Comorbidities of lifestyle-related diseases, such as diabetes mellitus (DM), chronic obstructive lung disease (COPD), hypertension (HT), hyperlipidemia (HL), chronic heart failure (CHF), insomnia, and cognitive impairment (C-I) were diagnosed and extracted by the physician who is specialists certified by the Japanese Society of Internal Medicine. Neuromuscular comorbidities such as musculoskeletal ambulation disability complex (MADS), rheumatoid arthritis (RA), osteoarthritis of the lower extremities (OA), joint contractures of the trunk or lower extremities (Contractures), disuse syndrome (Disuse), parkinsonism, and neuromuscular disorders (Parkinsonism) were also diagnosed and extracted by the physician who is specialists certified by the Japanese Orthopaedic Association and the Japanese College of Rheumatology.

The patient’s potential risk factors at baseline were extracted from several information resources. These are the patient’s T-score in the lumbar spine. The femoral neck was calculated from the DXA examination, from the medical records such as glucocorticoids (GCs), vitamin D (V-D), anti-osteoporotic drugs (OPD) administrations, and polypharmacy no less than seven kinds of tablet/capsules, as well as from blood test, such as estimated glomerular filtration rate with cystatin C (eGFR_CysC), albumin (ALB), calcium (Ca), procollagen type 1 amino-terminal propeptide (P1NP), tartrate-resistant acid phosphatase-5b (TRACP-5b), hemoglobin (Hgb), monocyte count (Mono), and lymphocyte count (Lymph).

## Statistical analyses

### 1. In the crude dataset

#### 1.1. Group comparison

The probability of incident clinical OF for each grade group using the SQ method classification at baseline was compared using an ANOVA Scheffe test. Our primary endpoint in this study is the development of incident vertebral fractures. As a secondary endpoint, the development of incident non-vertebral fracture (NVF) for each Grade group was also compared using the ANOVA Scheffe test. The mean value of the background clinical data for each Grade class was also compared using the ANOVA Scheffe tests.

Hazard ratios of the Grade groups for VF and NVF were compared using Kaplan-Meier curve analysis.

#### 1.2. Risk factor analysis

Cox regression analyses were examined in these patients. Development of incident clinical VF was set as a dependent factor (binary; yes/no). The time length of the follow-up period in months (continuous number) was set as survival time. Independent factors were patient’s sex (binary; male/female), age (continuous number), current smoke habitat (binary; yes/no), alcohol habitat (binary; yes/no), parent’s fracture (binary; yes/no), prevalent VF (binary; yes/no), prevalent NVF (binary; yes/no), positivity with VEC (binary; yes/no), presence of AAC (binary; yes/no), presence of each of comorbidities (binary; yes/no), T-score in the lumbar spine and femoral neck (LS and FN) (continuous number), drug factors such as GCs, V-D, OPD, polypharmacy (binary; yes/no), blood factors such as eGFR_CysC, ALB, Ca, P1NP, TRACP-5b, Hgb, and Mono (continuous number) were set as independent factors. Vertebral deformities classified with SQ at the baseline were set as an independent factor. This index was category data, but each grade was converted to an integer number for the statistical analysis.

First, these factors were examined with univariate models and then with a multivariate model for those that demonstrated a significant risk ratio. When the SQ is listed as a significant risk factor with the multivariate model, the study progresses to the next step. If not, the association between the significant factor and the SQ grade was evaluated.

#### 1.3. Differences in the variables among the SQ grade groups and the likelihood of the incident clinical VFs and NVFs in these variables analysis

We statistically compared variables among the SQ grade groups using Kruskal-Wallis ANOVA tests. Furthermore, these variables were analyzed for their likelihood of the incident clinical OFs separated into VFs and NVFs using Kaplan-Meier survival analysis. Before testing a Kaplan-Meier analysis, a receiver operating characteristic analysis (ROC) was performed to determine the cut-off index (COI) for continuous or stepwise numbers. We judged when a variable was statistically significant for both tests; the variable is the confounding factor for the incidence of VF and NVF.

### 2. In the dataset after propensity score matching (PSM)

#### 2.1 Group comparison after PSM

As a next step. the probability of incident VF or NVF for each SQ Grade group was compared using an ANOVA Scheffe test after the propensity of the confound factors in the crude dataset was leveled using the PSM technique. Furthermore, if the SQ grading does not listed for any of the incident osteoporotic fractures, the probability of the other incident fractures for each Grade group after PSM was also compared using the ANOVA Scheffe test. The mean value of the background clinical data for each Grade group was also compared using the ANOVA Scheffe tests. Hazard ratios of the Grade groups were also compared using Kaplan-Meier curve analysis.

#### 2.2 Risk factor analysis in the dataset after PSM

The dataset was examined using Cox regression analyses after the PSM procedures. The dependent factors were set as incident VFs and NVFs, and the independent factors were the same candidate risk factors used in the crude dataset. First, these factors were examined using univariate models, and then, for the factors that demonstrated a significant risk ratio, a multivariate model was used.

#### 3.1 Association between the significant factor and the SQ grade

An association between the significant factor and the SQ grade was examined using binary regression analysis when the factor was binary data or linear regression analysis when the factor was continuous data in the crude dataset. The probabilities of the incident VF and NVF for each SQ grade group were compared for each significant factor, which was separated by the COI using ROC analysis. The flowchart of the study, including inclusion and exclusion criteria, is shown in Figure 1.

**Figure 1:**
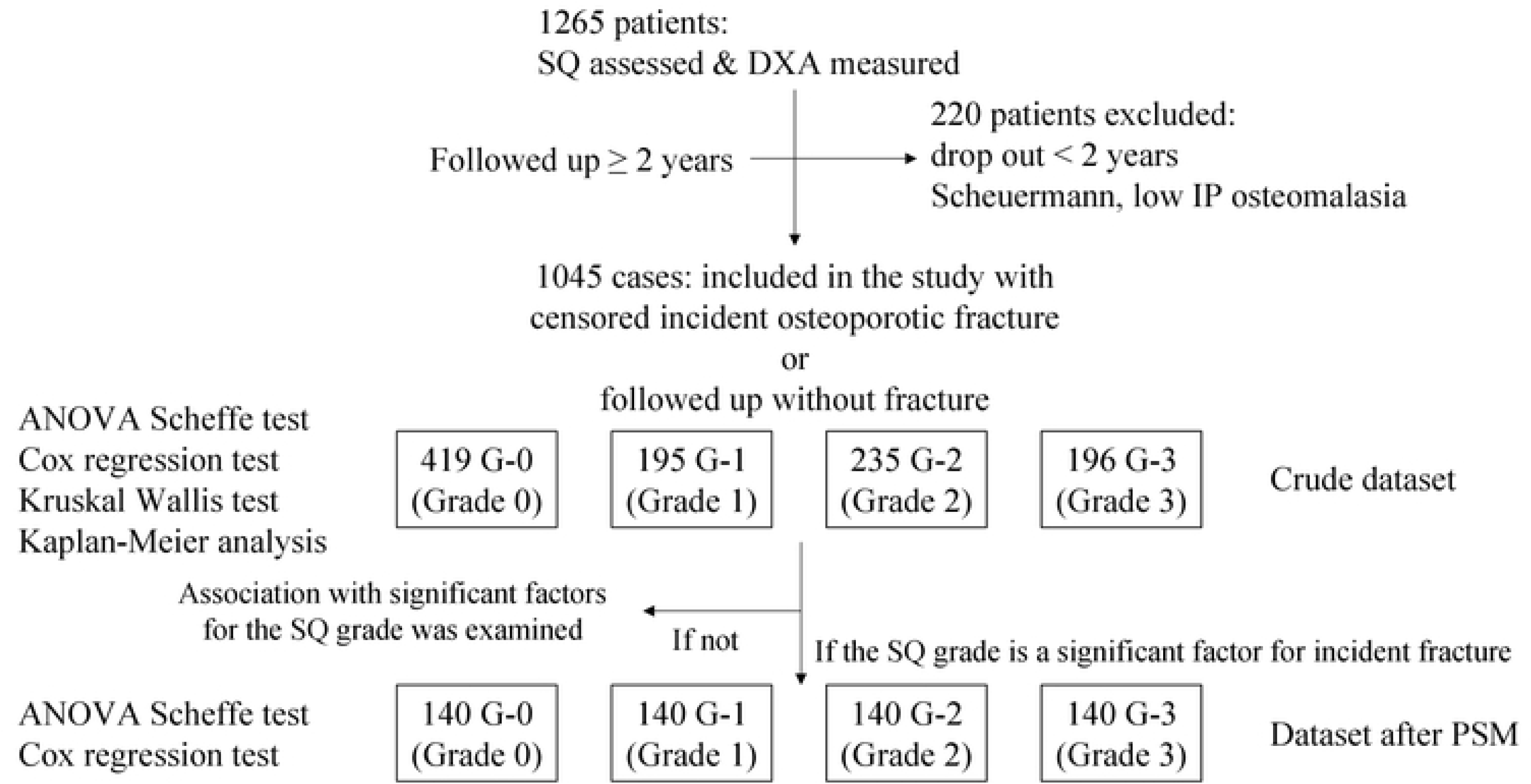
Flowchart of the study. Abbreviations: SQ, semi-quantitative method ; DXA, dual-energy X-ray absorptiometry ; PSM, propensity score matching

## Study Design Characteristics

Type of study: case-control study; STROBE checklist: written in another sheet; Primary outcome: development of an incident clinical vertebral fracture; Hypothesized effect size: 137. Secondary outcome: development of an incident clinical non-vertebral fracture; Hypothesized effect size: .

## Statistical procedures and software

All statistical procedures were performed using StatPlus:mac^®^ (AnalystSoft Inc., Walnut, CA, USA). Statistical significance was set below 5%.

## Ethical considerations

The study protocols and patient consent requirements were approved by the Ethics Committee of the associated institute (approval number: G-2023-3). The subjects and their families were informed that the personal information obtained in this study was anonymous and would only be used for analysis.

## Results

A total of 1045 patients, 143 male and 902 female, were included in the study. The mean age was 78.3 years old (SD: 10.8), and the mean follow-up period was 49.7 months (SD: 30.1, median: 33). In these patients, 419 in SQ G-0, 195 in SQ G-1, 235 in SQ G-2, and 196 in SQ G-3 were included in the crude dataset. The Cohen Kappa score was 0.865 (95% CI: 0.841 – 0.890).

### 1.1. Group comparison

A total of 59 patients had incident VFs in the crude dataset; in these, 13 (3.1%), 9 (4.6%), 13 (5.5%), and 24 (12.2%) in the G-0, G-1, G-2, and G-3 were included, respectively. G-3 had a significantly higher incident vertebral fracture rate than the other SQ groups (p<0.001).

There were 111 incident NVFs in the crude dataset; in these, 22 (5.3%), 25 (12.8%), 27 (11.5%), and 37 (18.9%) in the G-0, G-1, G-2, and G-3, respectively were included. The likelihood of incident VF in the crude dataset significantly differed among the SQ groups (p<0.001). Significant differences existed between the G-0 and G-3, the G-1 and G-3, and the G-2 and G-3 (p<0.001). In contrast, the likelihood of incident NVF in the crude dataset showed a significantly greater rate in the G-3 and G-1 than in the G-0 (Table 1).

**Table 1:**
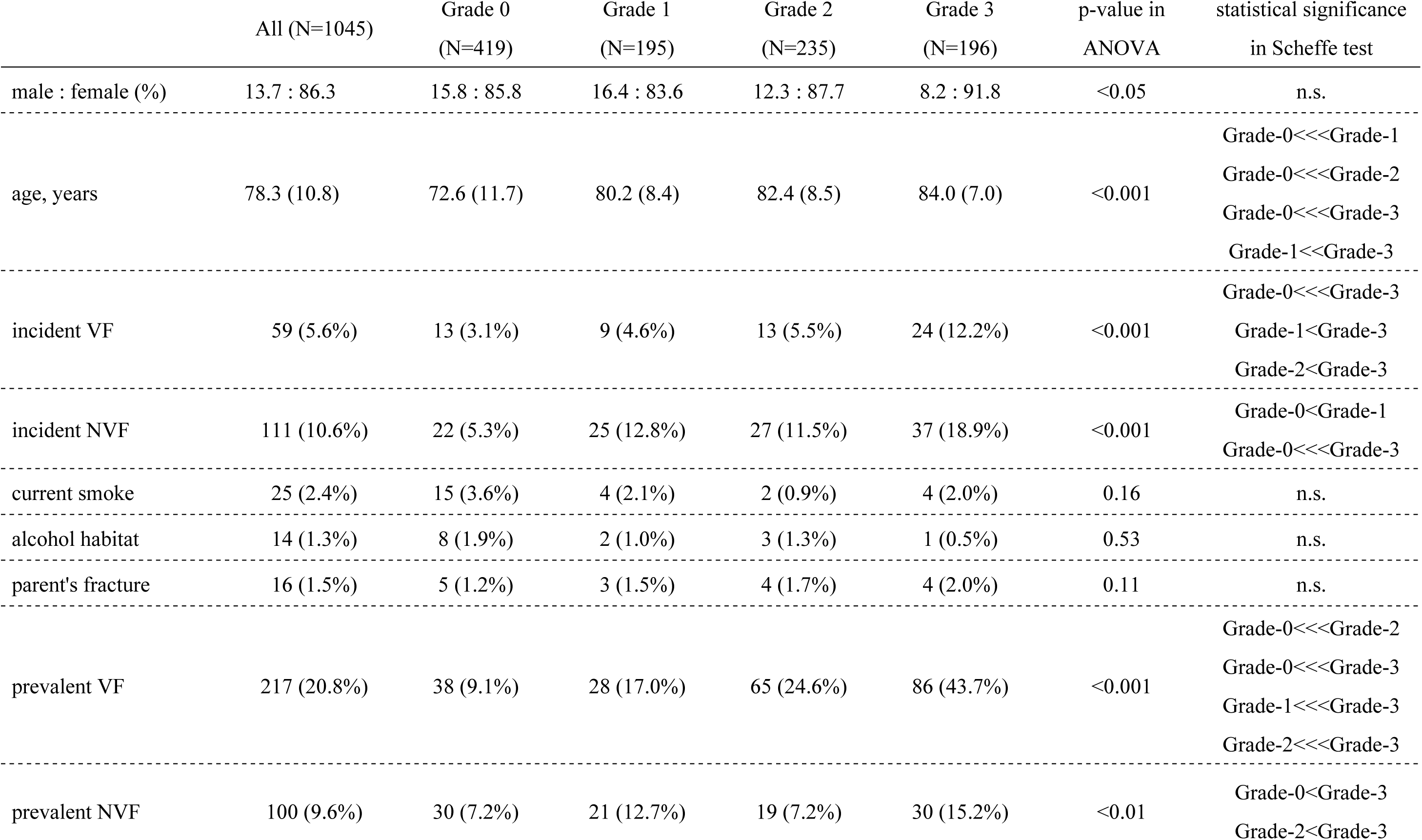
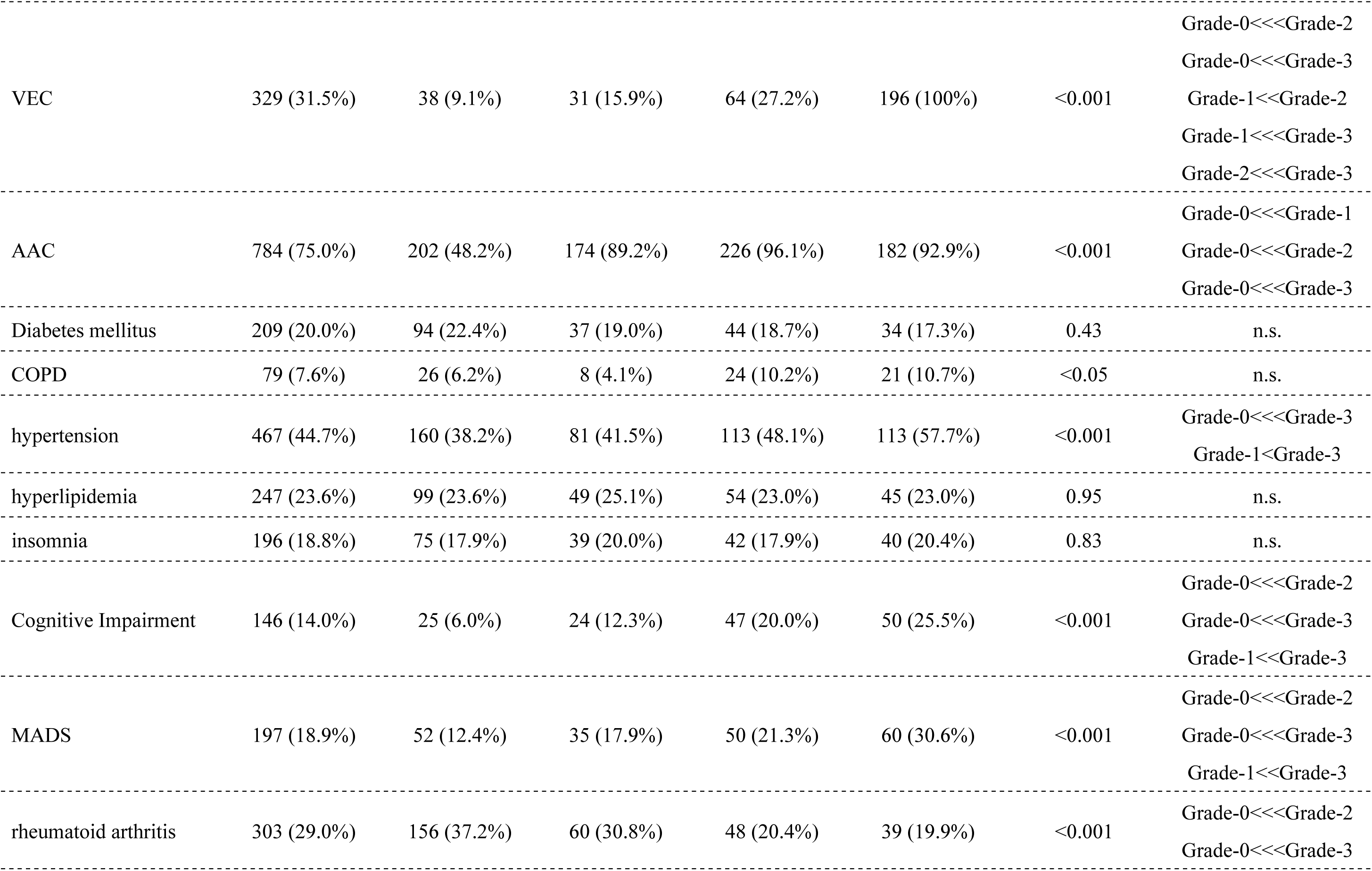
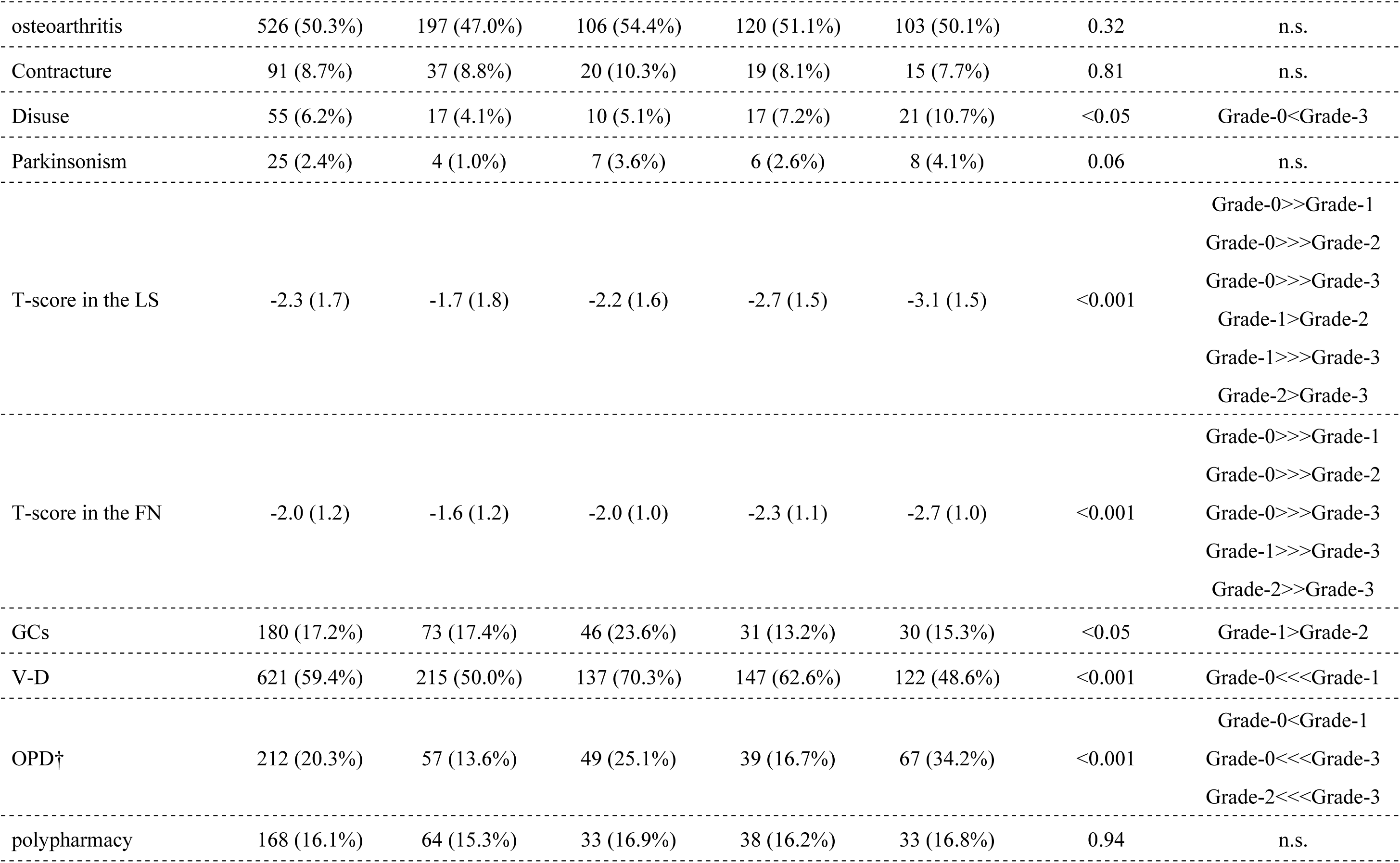
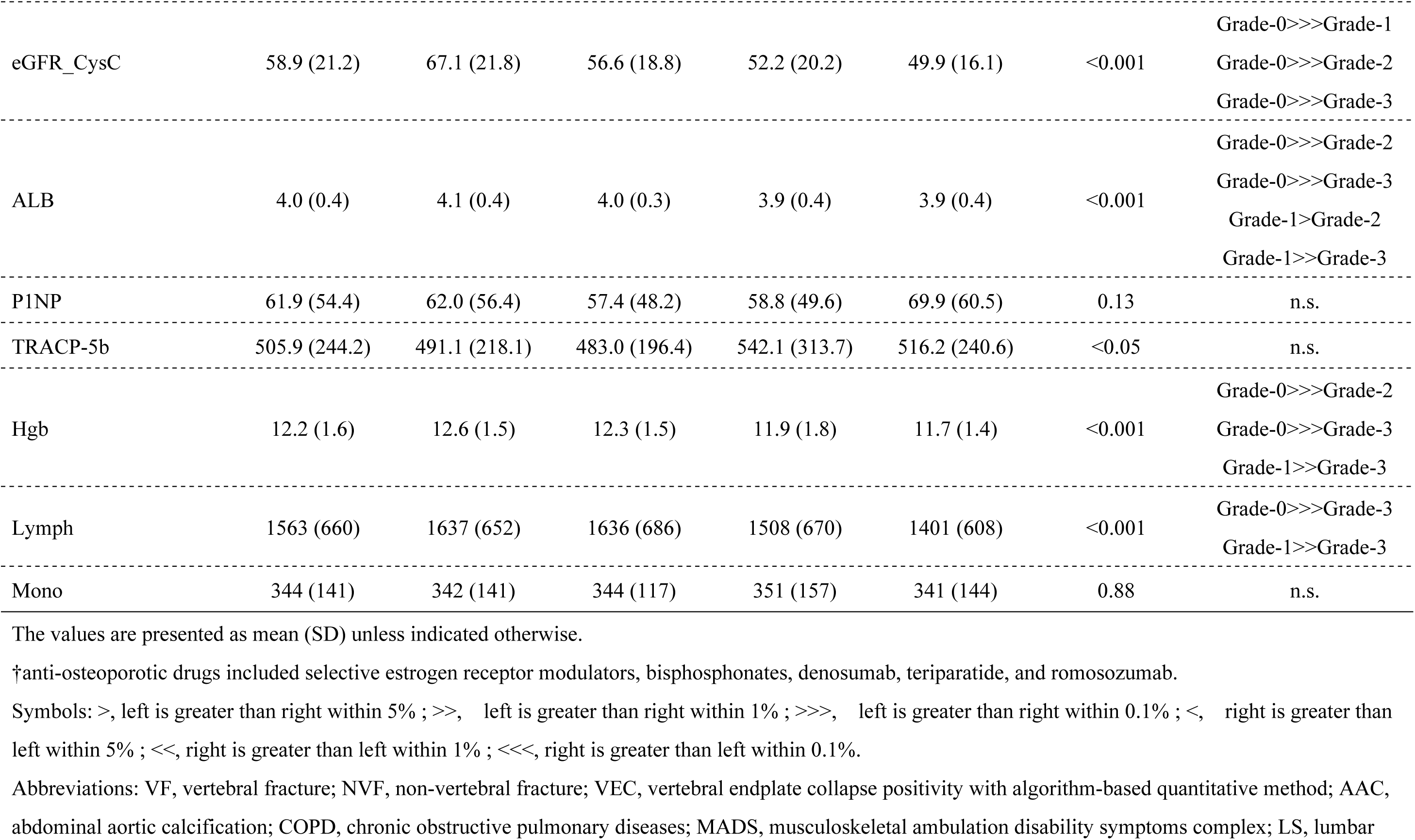
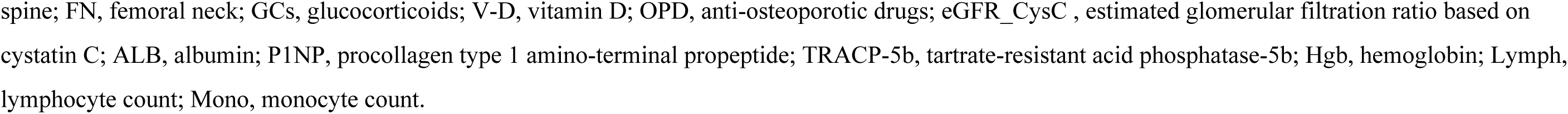
Demographic characteristics of the patients in the crude dataset.

Many significant factors were present when comparing the SQ groups. Factors that demonstrated significant differences among the SQ groups were mean age, follow-up length, prevalent VF and NVF, positivity with VEC, presence of AAC, HT, CHF, C-I, MADS, RA, and Disuse, mean T-score in the lumbar spine and the femoral neck, administration of GCs, V-D, and OPD at baseline, mean value of eGFR_CysC, mean serum ALB level, blood Hgb level, and Lymph level. More details are shown in Table 1. In Kaplan-Meier survival analysis, the Hazard ratios for incident VFs of G-2/G-0, G-3/G-0, and G-3/G-1 were significantly higher, with 2.13 (95%CI: 1.07 to 4.24), 4.47 (95%CI: 2.18 to 9.15), and 2.7 (95%CI: 1.19 to 6.44), respectively (Table 2).

**Table 2:** Hazard ratios of incident VFs in the crude and after PSM datasets.

| Groups | in the crude dataset |  | in the dataset after PSM |  |
| --- | --- | --- | --- | --- |
|  | HR (95%CI) | Std Err | HR (95%CI) | Std Err |
| G-0 / G-1 | 0.62 (0.30 – 1.26) | 0.36 | 1.14 (0.50 – 2.60) | 0.42 |
| G-0 / G-2 | **0.47 (0.24 – 0.93)** | 0.35 | 1.19 (0.51 – 2.75) | 0.43 |
| G-0 / G-3 | 0.22 (0.11 – 0.46) | 0.37 | 0.64 (0.27 – 1.47) | 0.43 |
| G-1 / G-0 | 1.62 (0.80 – 3.28) | 0.36 | 0.88 (0.38 – 2.00) | 0.42 |
| G-1 / G-2 | 0.76 (0.33 – 1.72) | 0.42 | 1.04 (0.44 – 2.45) | 0.44 |
| G-1 / G-3 | 0.36 (0.16 – 0.84) | 0.43 | 0.56 (0.24 – 1.31) | 0.44 |
| G-2 / G-0 | **2.13 (1.07 – 4.24)** | 0.35 | 0.84 (0.36 – 1.95) | 0.43 |
| G-2 / G-1 | 1.32 (0.58 – 3.00) | 0.42 | 0.96 (0.41 – 2.26) | 0.44 |
| G-2 / G-3 | 0.48 (0.21 – 1.09) | 0.42 | 0.53 (0.22 – 1.28) | 0.44 |
| G-3 / G-0 | **4.47 (2.18 – 9.15)** | 0.37 | 1.57 (0.68 – 3.64) | 0.43 |
| G-3 / G-1 | **2.76 (1.19 – 6.44)** | 0.43 | 1.80 (0.76 – 4.22) | 0.44 |
| G-3 / G-2 | 2.09 (0.91 – 4.79) | 0.42 | 1.87 (0.78 – 4.46) | 0.44 |

Bold style presents statistically significant.
Abbreviations: VF, vertebral fracture ; PSM, propensity score matching ; HR, Hazard ratio ; Std Err, Standard error ; 95%CI, 95% confident interval ; G-0, Grade 0 ; G-1, Grade 1 ; G-2, Grade 2 ; G-3, Grade 3

On the other hand, the Hazard ratios for incident NVFs of G-1/G-0, G-2/G-0, and G-3/G-0 were significantly higher, with 2.77 (95%CI: 1.68 to 4.59), 3.25 (95%CI: 1.94 to 5.46), and 5.03 (95%CI: 2.93 to 8.63), respectively. The other pairs had no significant Hazard ratios (Table 3).

**Table 3:** Hazard ratios of incident NVFs in the crude and after PSM datasets.

| Groups | in the crude dataset |  | in the dataset after PSM |  |
| --- | --- | --- | --- | --- |
|  | HR (95%CI) | Std Err | HR (95%CI) | Std Err |
| G-0 / G-1 | **0.36 (0.22 – 0.60)** | 0.26 | **0.34 (0.15 – 0.77)** | 0.42 |
| G-0 / G-2 | **0.31 (0.18 – 0.52)** | 0.26 | 0.65 (0.36 – 1.18) | 0.31 |
| G-0 / G-3 | **0.20 (0.12 – 0.34)** | 0.28 | **0.40 (0.20 – 0.79)** | 0.35 |
| G-1 / G-0 | **2.77 (1.68 – 4.59)** | 0.26 | **2.96 (1.29 – 6.79)** | 0.42 |
| G-1 / G-2 | 0.85 (0.46 – 1.57) | 0.31 | 1.93 (0.85 – 4.39) | 0.42 |
| G-1 / G-3 | 0.55 (0.29 – 1.04) | 0.32 | 1.19 (0.49 – 2.87) | 0.45 |
| G-2 / G-0 | **3.25 (1.94 – 5.46)** | 0.26 | 1.53 (0.85 – 2.78) | 0.30 |
| G-2 / G-1 | 1.17 (0.34 – 1.23) | 0.31 | 0.52 (0.23 – 1.18) | 0.42 |
| G-2 / G-3 | 0.65 (0.34 – 1.23) | 0.33 | 0.62 (0.32 – 1.20) | 0.34 |
| G-3 / G-0 | **5.03 (2.93 – 8.63)** | 0.28 | **2.49 (1.26 – 4.91)** | 0.35 |
| G-3 / G-1 | 1.81 (0.96 – 3.41) | 0.32 | 0.84 (0.35 – 2.04) | 0.45 |
| G-3 / G-2 | 1.55 (0.81 – 2.94) | 0.33 | 1.63 (0.83 – 3.17) | 0.34 |

Bold style presents statistically significant.
Abbreviations: NVF, non-vertebral fracture ; PSM, propensity score matching ; HR, Hazard ratio ; Std Err, Standard error ; 95%CI, 95% confident interval ; G-0, Grade 0 ; G-1, Grade 1 ; G-2, Grade 2 ; G-3, Grade 3

### 1.2. Risk factor analysis

In the crude dataset, older age, higher than Grade 1 in SQ, presence of AAC, CHF, C-I, MADS, lower T-score in the lumbar spine and the femoral neck, lower eGFR_CysC, lower Hgb, and lower Lymph had significantly higher risk ratios of incident VF using univariate models. Only those higher than Grade 1 in SQ had significantly higher risk ratios using multivariate models (Table 4).

**Table 4:**
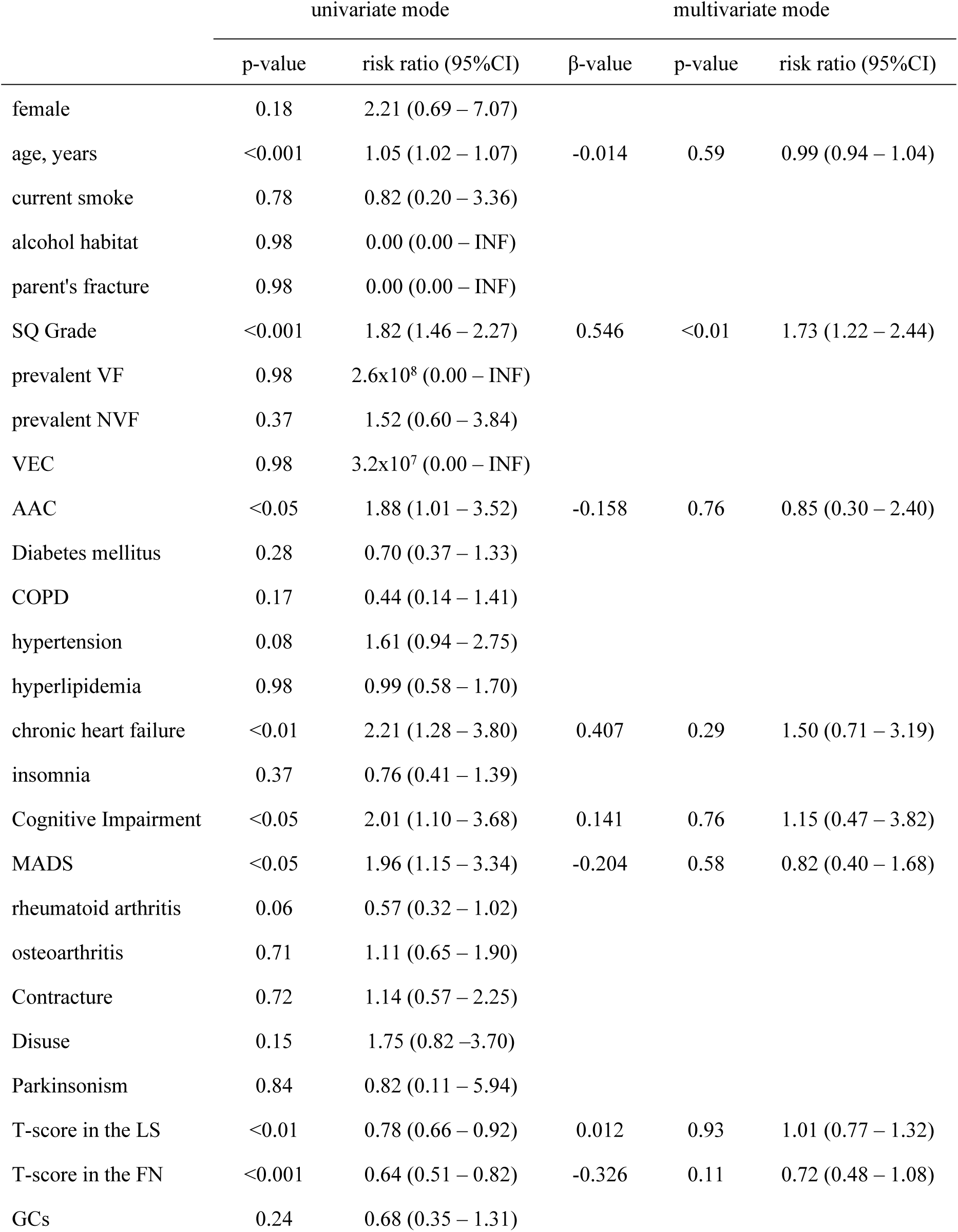
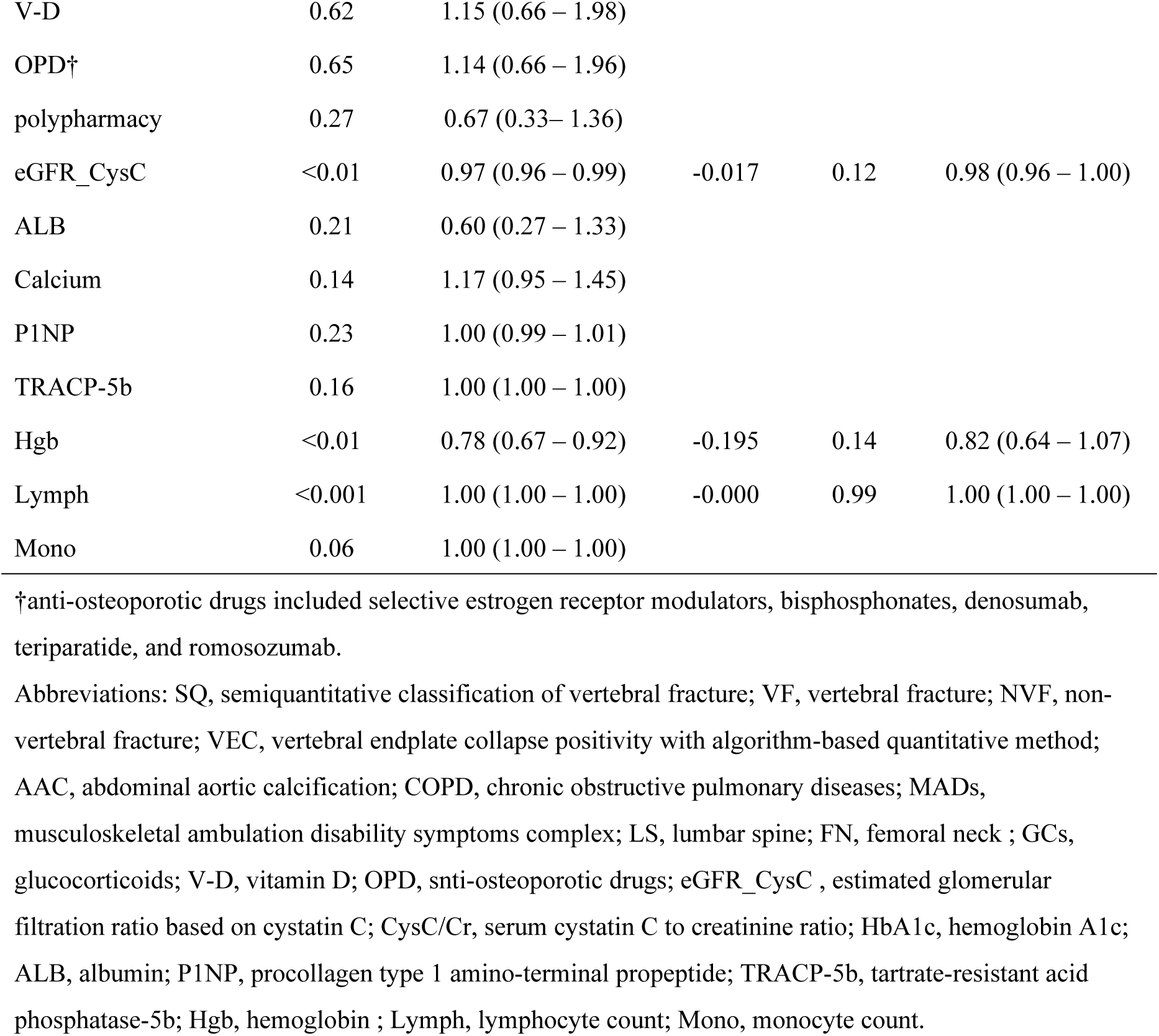
Results of a Cox regression analysis for incident VF development in the crude dataset.

|  | univariate mode |  | multivariate mode |  |  |
| --- | --- | --- | --- | --- | --- |
| | p-value | risk ratio (95%CI) | β-value | p-value | risk ratio (95%CI) |
| female | 0.18 | 2.21 (0.69 – 7.07) |  |  |  |
| age, years | <0.001 | 1.05 (1.02 – 1.07) | -0.014 | 0.59 | 0.99 (0.94 – 1.04) |
| current smoke | 0.78 | 0.82 (0.20 – 3.36) |  |  |  |
| alcohol habitat | 0.98 | 0.00 (0.00 – INF) |  |  |  |
| parent's fracture | 0.98 | 0.00 (0.00 – INF) |  |  |  |
| SQ Grade | <0.001 | 1.82 (1.46 – 2.27) | 0.546 | <0.01 | 1.73 (1.22 – 2.44) |
| prevalent VF | 0.98 | 2.6x10<sup>8</sup> (0.00 – INF) |  |  |  |
| prevalent NVF | 0.37 | 1.52 (0.60 – 3.84) |  |  |  |
| VEC | 0.98 | 3.2x10<sup>7</sup> (0.00 – INF) |  |  |  |
| AAC | <0.05 | 1.88 (1.01 – 3.52) | -0.158 | 0.76 | 0.85 (0.30 – 2.40) |
| Diabetes mellitus | 0.28 | 0.70 (0.37 – 1.33) |  |  |  |
| COPD | 0.17 | 0.44 (0.14 – 1.41) |  |  |  |
| hypertension | 0.08 | 1.61 (0.94 – 2.75) |  |  |  |
| hyperlipidemia | 0.98 | 0.99 (0.58 – 1.70) |  |  |  |
| chronic heart failure | <0.01 | 2.21 (1.28 – 3.80) | 0.407 | 0.29 | 1.50 (0.71 – 3.19) |
| insomnia | 0.37 | 0.76 (0.41 – 1.39) |  |  |  |
| Cognitive Impairment | <0.05 | 2.01 (1.10 – 3.68) | 0.141 | 0.76 | 1.15 (0.47 – 3.82) |
| MADS | <0.05 | 1.96 (1.15 – 3.34) | -0.204 | 0.58 | 0.82 (0.40 – 1.68) |
| rheumatoid arthritis | 0.06 | 0.57 (0.32 – 1.02) |  |  |  |
| osteoarthritis | 0.71 | 1.11 (0.65 – 1.90) |  |  |  |
| Contracture | 0.72 | 1.14 (0.57 – 2.25) |  |  |  |
| Disuse | 0.15 | 1.75 (0.82 – 3.70) |  |  |  |
| Parkinsonism | 0.84 | 0.82 (0.11 – 5.94) |  |  |  |
| T-score in the LS | <0.01 | 0.78 (0.66 – 0.92) | 0.012 | 0.93 | 1.01 (0.77 – 1.32) |
| T-score in the FN | <0.001 | 0.64 (0.51 – 0.82) | -0.326 | 0.11 | 0.72 (0.48 – 1.08) |
| GCs | 0.24 | 0.68 (0.35 – 1.31) |  |  |  |
| V-D | 0.62 | 1.15 (0.66 – 1.98) |  |  |  |
| OPD† | 0.65 | 1.14 (0.66 – 1.96) |  |  |  |
| polypharmacy | 0.27 | 0.67 (0.33– 1.36) |  |  |  |
| eGFR_CysC | <0.01 | 0.97 (0.96 – 0.99) | -0.017 | 0.12 | 0.98 (0.96 – 1.00) |
| ALB | 0.21 | 0.60 (0.27 – 1.33) |  |  |  |
| Calcium | 0.14 | 1.17 (0.95 – 1.45) |  |  |  |
| P1NP | 0.23 | 1.00 (0.99 – 1.01) |  |  |  |
| TRACP-5b | 0.16 | 1.00 (1.00 – 1.00) |  |  |  |
| Hgb | <0.01 | 0.78 (0.67 – 0.92) | -0.195 | 0.14 | 0.82 (0.64 – 1.07) |
| Lymph | <0.001 | 1.00 (1.00 – 1.00) | -0.000 | 0.99 | 1.00 (1.00 – 1.00) |
| Mono | 0.06 | 1.00 (1.00 – 1.00) |  |  |  |

†anti-osteoporotic drugs included selective estrogen receptor modulators, bisphosphonates, denosumab, teriparatide, and romosozumab.
Abbreviations: SQ, semiquantitative classification of vertebral fracture; VF, vertebral fracture; NVF, non-vertebral fracture; VEC, vertebral endplate collapse positivity with algorithm-based quantitative method; AAC, abdominal aortic calcification; COPD, chronic obstructive pulmonary diseases; MADs, musculoskeletal ambulation disability symptoms complex; LS, lumbar spine; FN, femoral neck ; GCs, glucocorticoids; V-D, vitamin D; OPD, anti-osteoporotic drugs; eGFR\_CysC , estimated glomerular filtration ratio based on cystatin C; CysC/Cr, serum cystatin C to creatinine ratio; HbA1c, hemoglobin A1c; ALB, albumin; P1NP, procollagen type 1 amino-terminal propeptide; TRACP-5b, tartrate-resistant acid phosphatase-5b; Hgb, hemoglobin ; Lymph, lymphocyte count; Mono, monocyte count.

On the other hand, in addition to those, the presence of prevalent VF, NVF, VEC, HT, Disuse, Parkinsonism, lower P1NP, lower ALB, lower Hgb, lower Lymph, and lower Mono significantly increased the risk ratio of incident NVF using univariate models. Only the presence of HT significantly increased the risk ratio using a multivariate model (Table 5).

**Table 5:**
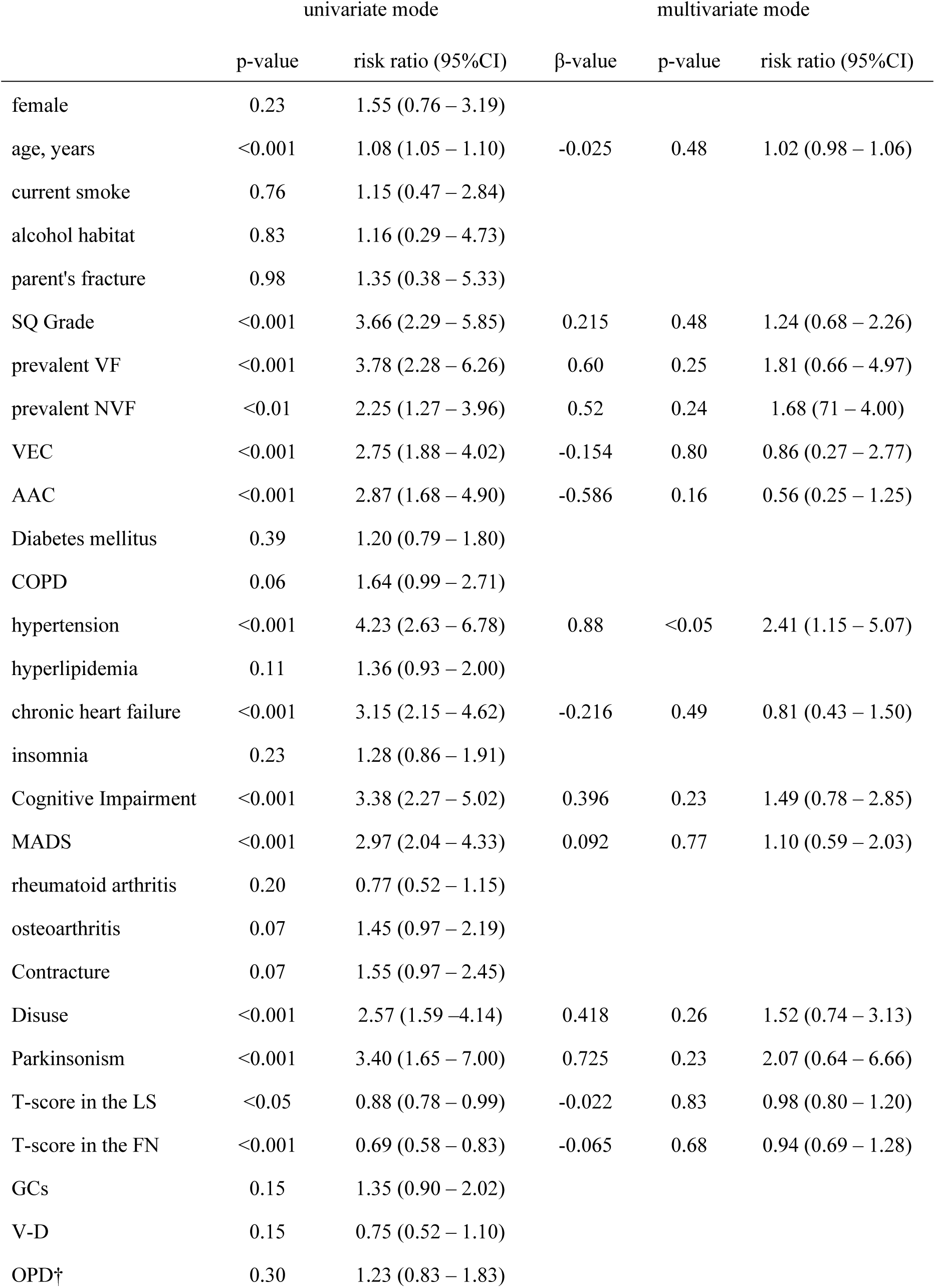
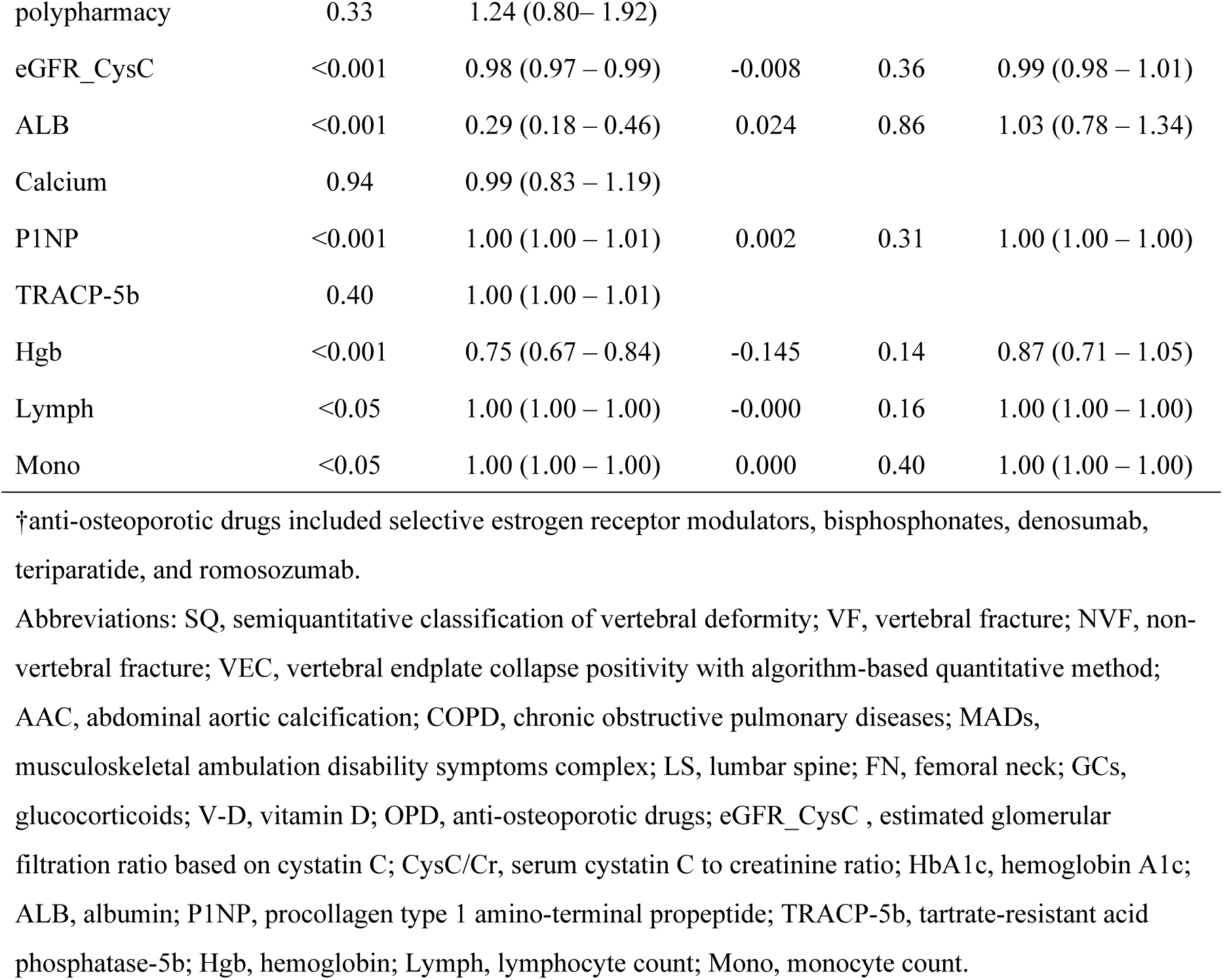
Results of a Cox regression analysis for incident NVF development in the crude dataset.

| | univariate mode | | β-value | multivariate mode | |
| --- | --- | --- | --- | --- | --- |
|  | p-value | risk ratio (95%CI) |  | p-value | risk ratio (95%CI) |
| female | 0.23 | 1.55 (0.76 – 3.19) |  |  |  |
| age, years | <0.001 | 1.08 (1.05 – 1.10) | -0.025 | 0.48 | 1.02 (0.98 – 1.06) |
| current smoke | 0.76 | 1.15 (0.47 – 2.84) |  |  |  |
| alcohol habitat | 0.83 | 1.16 (0.29 – 4.73) |  |  |  |
| parent's fracture | 0.98 | 1.35 (0.38 – 5.33) |  |  |  |
| SQ Grade | <0.001 | 3.66 (2.29 – 5.85) | 0.215 | 0.48 | 1.24 (0.68 – 2.26) |
| prevalent VF | <0.001 | 3.78 (2.28 – 6.26) | 0.60 | 0.25 | 1.81 (0.66 – 4.97) |
| prevalent NVF | <0.01 | 2.25 (1.27 – 3.96) | 0.52 | 0.24 | 1.68 (0.71 – 4.00) |
| VEC | <0.001 | 2.75 (1.88 – 4.02) | -0.154 | 0.80 | 0.86 (0.27 – 2.77) |
| AAC | <0.001 | 2.87 (1.68 – 4.90) | -0.586 | 0.16 | 0.56 (0.25 – 1.25) |
| Diabetes mellitus | 0.39 | 1.20 (0.79 – 1.80) |  |  |  |
| COPD | 0.06 | 1.64 (0.99 – 2.71) |  |  |  |
| hypertension | <0.001 | 4.23 (2.63 – 6.78) | 0.88 | <0.05 | 2.41 (1.15 – 5.07) |
| hyperlipidemia | 0.11 | 1.36 (0.93 – 2.00) |  |  |  |
| chronic heart failure | <0.001 | 3.15 (2.15 – 4.62) | -0.216 | 0.49 | 0.81 (0.43 – 1.50) |
| insomnia | 0.23 | 1.28 (0.86 – 1.91) |  |  |  |
| Cognitive Impairment | <0.001 | 3.38 (2.27 – 5.02) | 0.396 | 0.23 | 1.49 (0.78 – 2.85) |
| MADS | <0.001 | 2.97 (2.04 – 4.33) | 0.092 | 0.77 | 1.10 (0.59 – 2.03) |
| rheumatoid arthritis | 0.20 | 0.77 (0.52 – 1.15) |  |  |  |
| osteoarthritis | 0.07 | 1.45 (0.97 – 2.19) |  |  |  |
| Contracture | 0.07 | 1.55 (0.97 – 2.45) |  |  |  |
| Disuse | <0.001 | 2.57 (1.59 – 4.14) | 0.418 | 0.26 | 1.52 (0.74 – 3.13) |
| Parkinsonism | <0.001 | 3.40 (1.65 – 7.00) | 0.725 | 0.23 | 2.07 (0.64 – 6.66) |
| T-score in the LS | <0.05 | 0.88 (0.78 – 0.99) | -0.022 | 0.83 | 0.98 (0.80 – 1.20) |
| T-score in the FN | <0.001 | 0.69 (0.58 – 0.83) | -0.065 | 0.68 | 0.94 (0.69 – 1.28) |
| GCs | 0.15 | 1.35 (0.90 – 2.02) |  |  |  |
| V-D | 0.15 | 0.75 (0.52 – 1.10) |  |  |  |
| OPD† | 0.30 | 1.23 (0.83 – 1.83) |  |  |  |
| polypharmacy | 0.33 | 1.24 (0.80– 1.92) |  |  |  |
| eGFR_CysC | <0.001 | 0.98 (0.97 – 0.99) | -0.008 | 0.36 | 0.99 (0.98 – 1.01) |
| ALB | <0.001 | 0.29 (0.18 – 0.46) | 0.024 | 0.86 | 1.03 (0.78 – 1.34) |
| Calcium | 0.94 | 0.99 (0.83 – 1.19) |  |  |  |
| P1NP | <0.001 | 1.00 (1.00 – 1.01) | 0.002 | 0.31 | 1.00 (1.00 – 1.00) |
| TRACP-5b | 0.40 | 1.00 (1.00 – 1.01) |  |  |  |
| Hgb | <0.001 | 0.75 (0.67 – 0.84) | -0.145 | 0.14 | 0.87 (0.71 – 1.05) |
| Lymph | <0.05 | 1.00 (1.00 – 1.00) | -0.000 | 0.16 | 1.00 (1.00 – 1.00) |
| Mono | <0.05 | 1.00 (1.00 – 1.00) | 0.000 | 0.40 | 1.00 (1.00 – 1.00) |

†anti-osteoporotic drugs included selective estrogen receptor modulators, bisphosphonates, denosumab, teriparatide, and romosozumab.
Abbreviations: SQ, semiquantitative classification of vertebral deformity; VF, vertebral fracture; NVF, non-vertebral fracture; VEC, vertebral endplate collapse positivity with algorithm-based quantitative method; AAC, abdominal aortic calcification; COPD, chronic obstructive pulmonary diseases; MADs, musculoskeletal ambulation disability symptoms complex; LS, lumbar spine; FN, femoral neck; GCs, glucocorticoids; V-D, vitamin D; OPD, anti-osteoporotic drugs; eGFR\_CysC , estimated glomerular filtration ratio based on cystatin C; CysC/Cr, serum cystatin C to creatinine ratio; HbA1c, hemoglobin A1c; ALB, albumin; P1NP, procollagen type 1 amino-terminal propeptide; TRACP-5b, tartrate-resistant acid phosphatase-5b; Hgb, hemoglobin; Lymph, lymphocyte count; Mono, monocyte count.

### 1.3. Differences in the variables among the SQ grade groups and the likelihood of the incident VFs and NVFs in these variables analysis

Age at baseline, presence of prevalent VF, presence of VEC using ABQ, presence of AAC, HT, CHF, C-I, MADS, RA, SpA, T-score in the lumbar spine and femoral neck, administration of Vitamin-D and OPD, eGFR_CysC, serum ALB level, blood HGB level, and Lymph count were significantly different among the SQ grade groups using a Kruskal-Wallis test. In contrast, the likelihood of the incident VFs in these variables showed statistical significance in VEC with ABQ, presence of CHF, C-I, MADS, T-score in the lumbar spine, and eGFR_CysC using Kaplan-Meier survival analysis. Thus, these variables were judged as the confounding factors of the SQ grade for the incident VFs. On the other hand, the likelihood of the incident NVFs in these variables showed statistical significance in the presence of prevalent VF, VEC with ABQ, AAC, HT, CHF, C-I, MADS, T-score in the LS and FN, eGFR_CysC, ALB, Hgb, and Lymph using Kaplan-Meier survival analysis (Table 6).

**Table 6:**
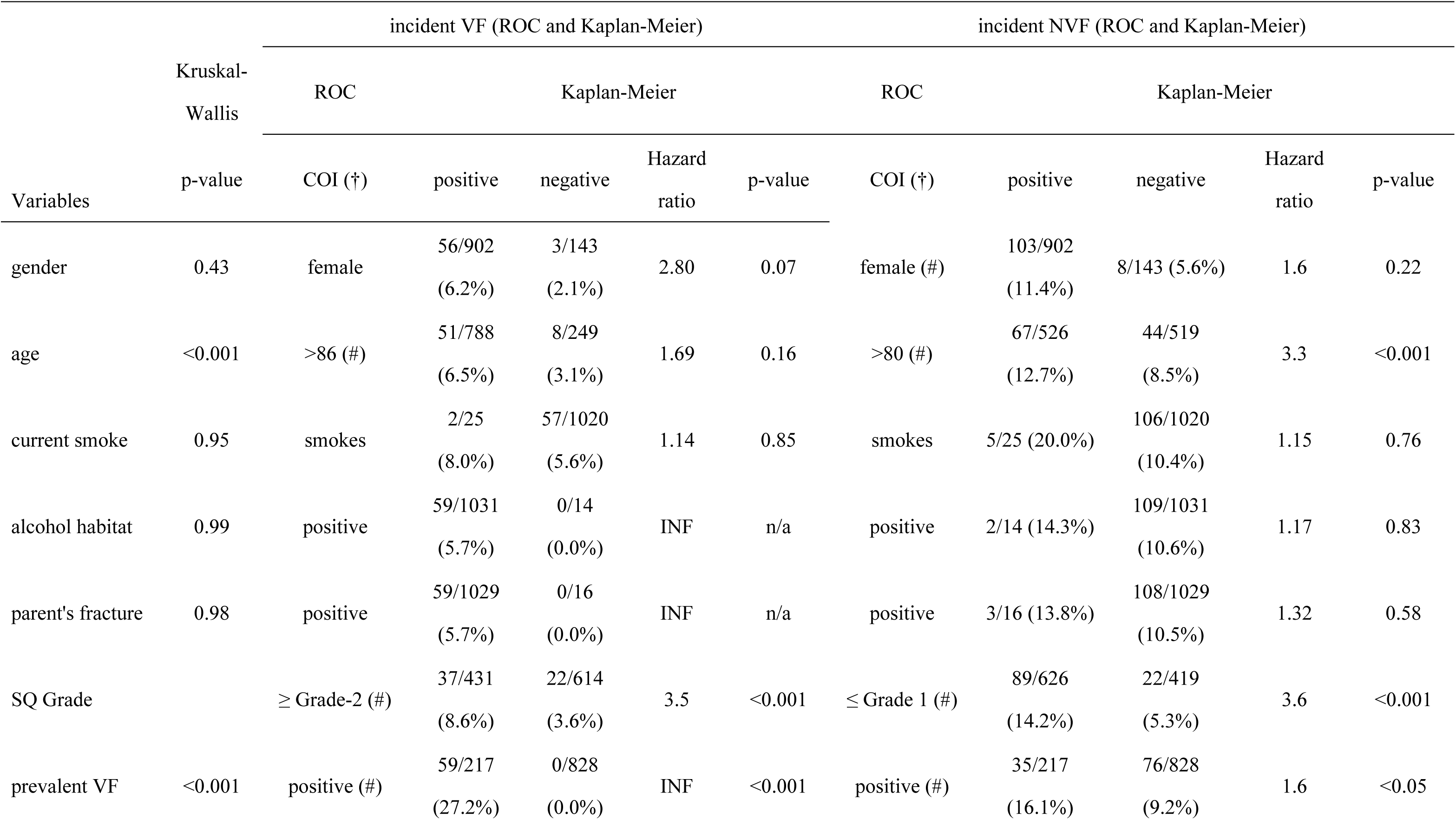
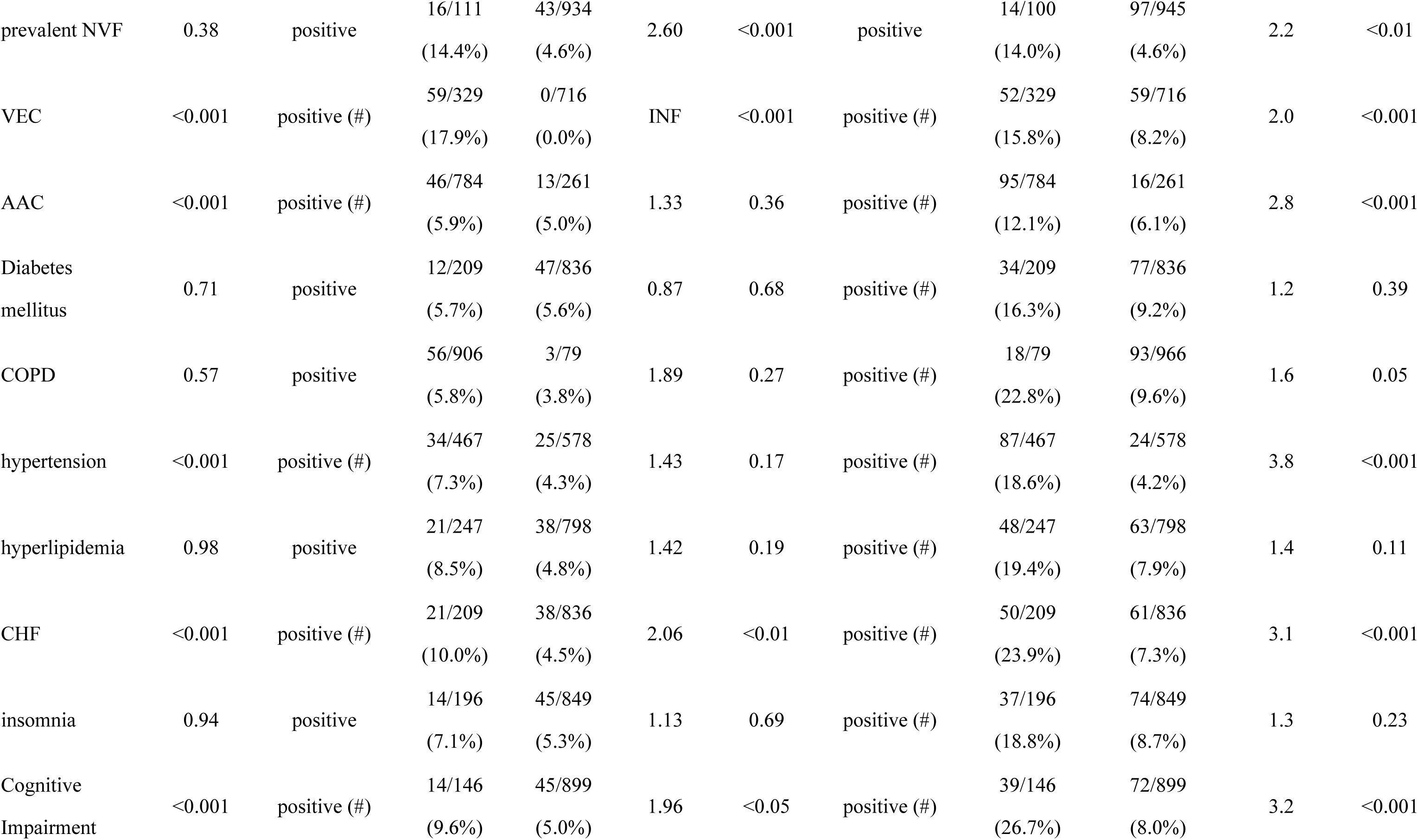
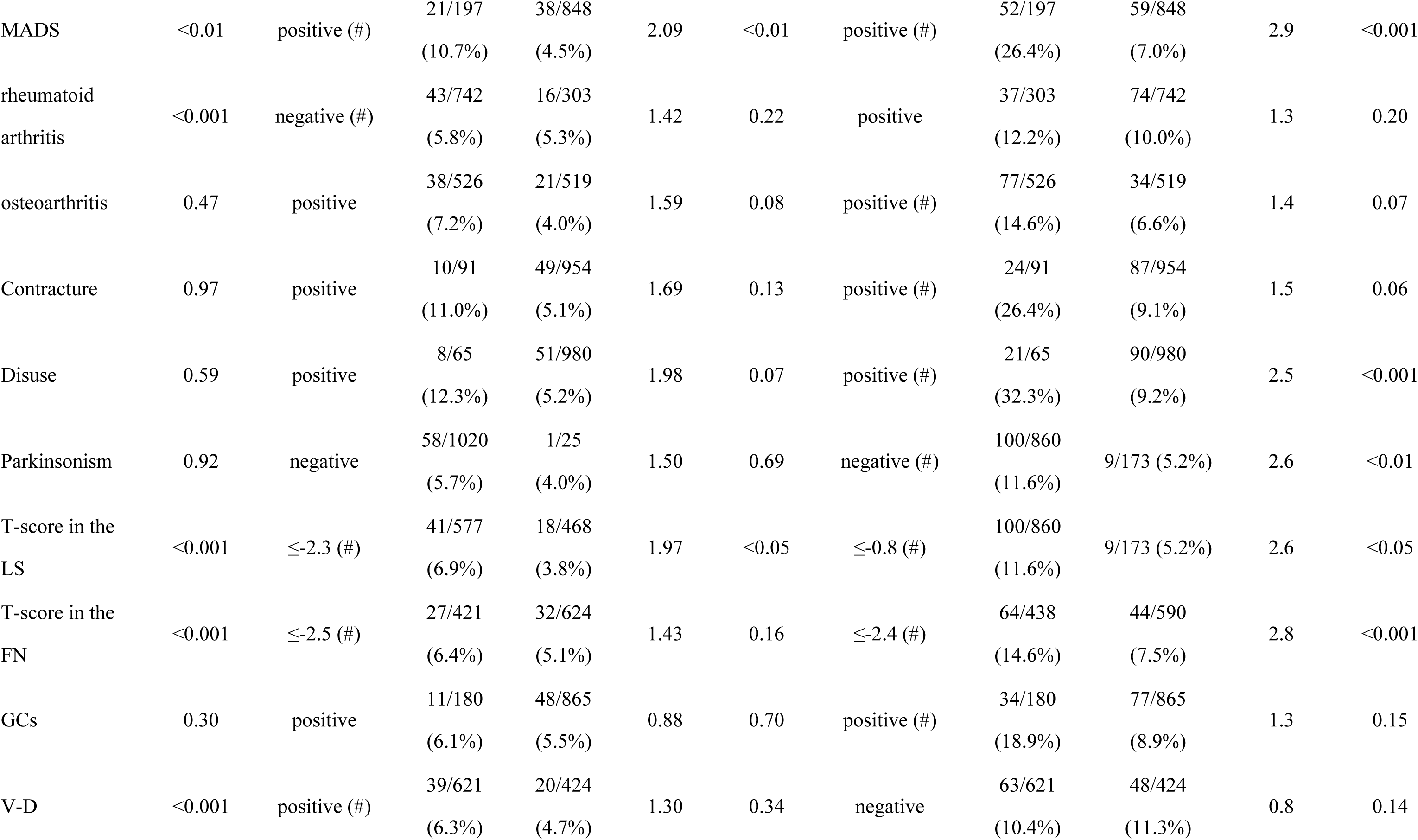
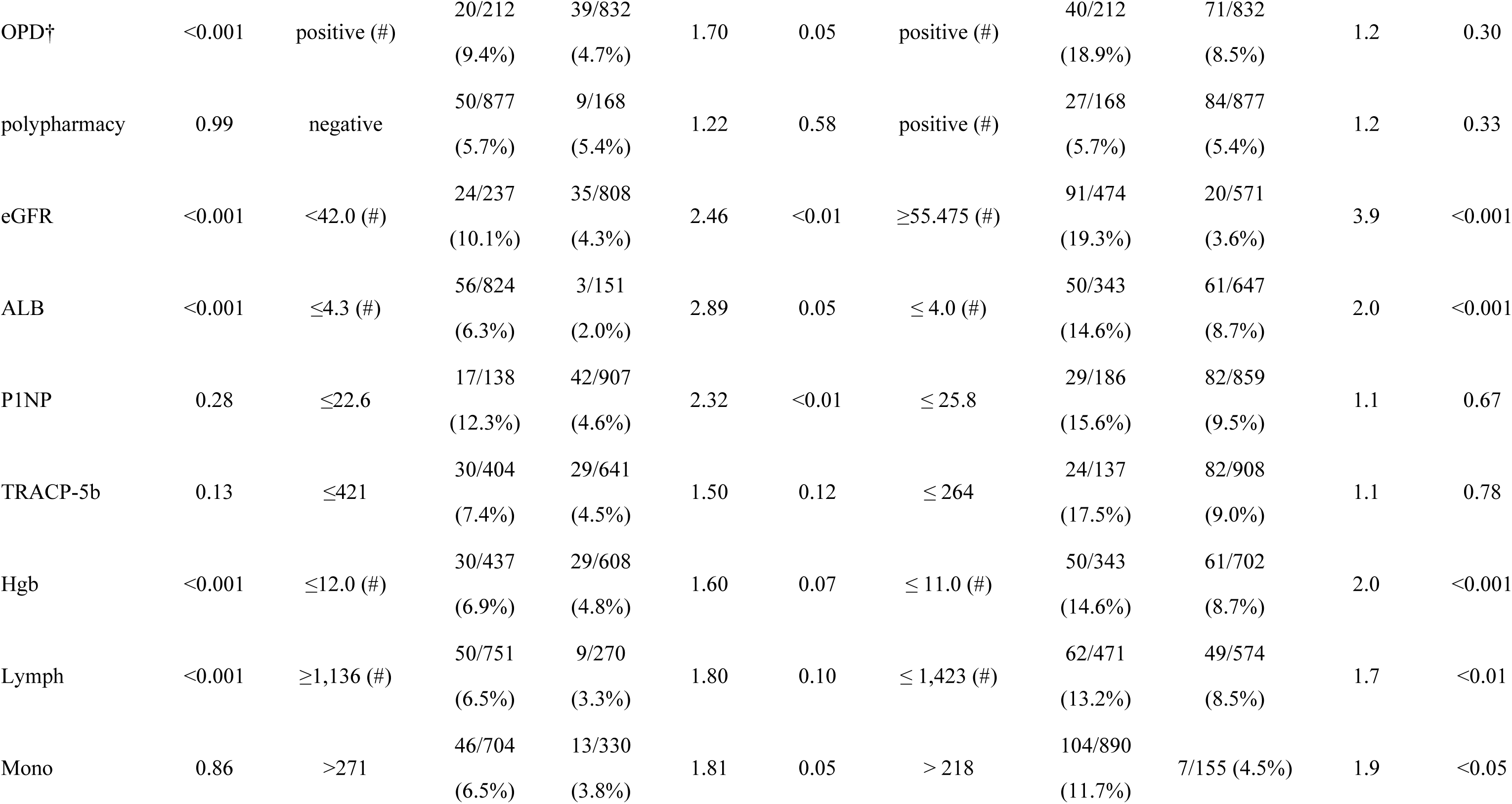
Statistical differences in variables among SQ Grades examined with Kruskal-Wallis ANOVA test presented with *p*-values and likelihood of incident VFs and NVFs of the variables examined with a receiver operating characteristic (ROC) and Kaplan-Meier analyses presented with cut-off index (COI), Hazard ratio, and p-values

| Variables | Kruskal-Wallis<br>p-value | incident VF (ROC and Kaplan-Meier) |  |  |  |  | incident NVF (ROC and Kaplan-Meier) |  |  |  |  |
| --- | --- | --- | --- | --- | --- | --- | --- | --- | --- | --- | --- |
|  |  | ROC | Kaplan-Meier |  |  | p-value | ROC | Kaplan-Meier |  |  | p-value |
|  |  | COI (†) | positive | negative | Hazard ratio |  | COI (†) | positive | negative | Hazard ratio |  |
| gender | 0.43 | female | 56/902<br>(6.2%) | 3/143<br>(2.1%) | 2.80 | 0.07 | female (#) | 103/902<br>(11.4%) | 8/143 (5.6%) | 1.6 | 0.22 |
| age | <0.001 | >86 (#) | 51/788<br>(6.5%) | 8/249<br>(3.1%) | 1.69 | 0.16 | >80 (#) | 67/526<br>(12.7%) | 44/519<br>(8.5%) | 3.3 | <0.001 |
| current smoke | 0.95 | smokes | 2/25<br>(8.0%) | 57/1020<br>(5.6%) | 1.14 | 0.85 | smokes | 5/25 (20.0%) | 106/1020<br>(10.4%) | 1.15 | 0.76 |
| alcohol habitat | 0.99 | positive | 59/1031<br>(5.7%) | 0/14<br>(0.0%) | INF | n/a | positive | 2/14 (14.3%) | 109/1031<br>(10.6%) | 1.17 | 0.83 |
| parent's fracture | 0.98 | positive | 59/1029<br>(5.7%) | 0/16<br>(0.0%) | INF | n/a | positive | 3/16 (13.8%) | 108/1029<br>(10.5%) | 1.32 | 0.58 |
| SQ Grade |  | ≥ Grade-2 (#) | 37/431<br>(8.6%) | 22/614<br>(3.6%) | 3.5 | <0.001 | ≤ Grade 1 (#) | 89/626<br>(14.2%) | 22/419<br>(5.3%) | 3.6 | <0.001 |
| prevalent VF | <0.001 | positive (#) | 59/217<br>(27.2%) | 0/828<br>(0.0%) | INF | <0.001 | positive (#) | 35/217<br>(16.1%) | 76/828<br>(9.2%) | 1.6 | <0.05 |
| prevalent NVF | 0.38 | positive | 16/111<br>(14.4%) | 43/934<br>(4.6%) | 2.60 | <0.001 | positive | 14/100<br>(14.0%) | 97/945<br>(4.6%) | 2.2 | <0.01 |
| VEC | <0.001 | positive (#) | 59/329<br>(17.9%) | 0/716<br>(0.0%) | INF | <0.001 | positive (#) | 52/329<br>(15.8%) | 59/716<br>(8.2%) | 2.0 | <0.001 |
| AAC | <0.001 | positive (#) | 46/784<br>(5.9%) | 13/261<br>(5.0%) | 1.33 | 0.36 | positive (#) | 95/784<br>(12.1%) | 16/261<br>(6.1%) | 2.8 | <0.001 |
| Diabetes<br>mellitus | 0.71 | positive | 12/209<br>(5.7%) | 47/836<br>(5.6%) | 0.87 | 0.68 | positive (#) | 34/209<br>(16.3%) | 77/836<br>(9.2%) | 1.2 | 0.39 |
| COPD | 0.57 | positive | 56/906<br>(5.8%) | 3/79<br>(3.8%) | 1.89 | 0.27 | positive (#) | 18/79<br>(22.8%) | 93/966<br>(9.6%) | 1.6 | 0.05 |
| hypertension | <0.001 | positive (#) | 34/467<br>(7.3%) | 25/578<br>(4.3%) | 1.43 | 0.17 | positive (#) | 87/467<br>(18.6%) | 24/578<br>(4.2%) | 3.8 | <0.001 |
| hyperlipidemia | 0.98 | positive | 21/247<br>(8.5%) | 38/798<br>(4.8%) | 1.42 | 0.19 | positive (#) | 48/247<br>(19.4%) | 63/798<br>(7.9%) | 1.4 | 0.11 |
| CHF | <0.001 | positive (#) | 21/209<br>(10.0%) | 38/836<br>(4.5%) | 2.06 | <0.01 | positive (#) | 50/209<br>(23.9%) | 61/836<br>(7.3%) | 3.1 | <0.001 |
| insomnia | 0.94 | positive | 14/196<br>(7.1%) | 45/849<br>(5.3%) | 1.13 | 0.69 | positive (#) | 37/196<br>(18.8%) | 74/849<br>(8.7%) | 1.3 | 0.23 |
| Cognitive<br>Impairment | <0.001 | positive (#) | 14/146<br>(9.6%) | 45/899<br>(5.0%) | 1.96 | <0.05 | positive (#) | 39/146<br>(26.7%) | 72/899<br>(8.0%) | 3.2 | <0.001 |
| MADS | <0.01 | positive (#) | 21/197<br>(10.7%) | 38/848<br>(4.5%) | 2.09 | <0.01 | positive (#) | 52/197<br>(26.4%) | 59/848<br>(7.0%) | 2.9 | <0.001 |
| rheumatoid<br>arthritis | <0.001 | negative (#) | 43/742<br>(5.8%) | 16/303<br>(5.3%) | 1.42 | 0.22 | positive | 37/303<br>(12.2%) | 74/742<br>(10.0%) | 1.3 | 0.20 |
| osteoarthritis | 0.47 | positive | 38/526<br>(7.2%) | 21/519<br>(4.0%) | 1.59 | 0.08 | positive (#) | 77/526<br>(14.6%) | 34/519<br>(6.6%) | 1.4 | 0.07 |
| Contracture | 0.97 | positive | 10/91<br>(11.0%) | 49/954<br>(5.1%) | 1.69 | 0.13 | positive (#) | 24/91<br>(26.4%) | 87/954<br>(9.1%) | 1.5 | 0.06 |
| Disuse | 0.59 | positive | 8/65<br>(12.3%) | 51/980<br>(5.2%) | 1.98 | 0.07 | positive (#) | 21/65<br>(32.3%) | 90/980<br>(9.2%) | 2.5 | <0.001 |
| Parkinsonism | 0.92 | negative | 58/1020<br>(5.7%) | 1/25<br>(4.0%) | 1.50 | 0.69 | negative (#) | 100/860<br>(11.6%) | 9/173 (5.2%) | 2.6 | <0.01 |
| T-score in the<br>LS | <0.001 | ≤-2.3 (#) | 41/577<br>(6.9%) | 18/468<br>(3.8%) | 1.97 | <0.05 | ≤-0.8 (#) | 100/860<br>(11.6%) | 9/173 (5.2%) | 2.6 | <0.05 |
| T-score in the<br>FN | <0.001 | ≤-2.5 (#) | 27/421<br>(6.4%) | 32/624<br>(5.1%) | 1.43 | 0.16 | ≤-2.4 (#) | 64/438<br>(14.6%) | 44/590<br>(7.5%) | 2.8 | <0.001 |
| GCs | 0.30 | positive | 11/180<br>(6.1%) | 48/865<br>(5.5%) | 0.88 | 0.70 | positive (#) | 34/180<br>(18.9%) | 77/865<br>(8.9%) | 1.3 | 0.15 |
| V-D | <0.001 | positive (#) | 39/621<br>(6.3%) | 20/424<br>(4.7%) | 1.30 | 0.34 | negative | 63/621<br>(10.4%) | 48/424<br>(11.3%) | 0.8 | 0.14 |
| OPD† | <0.001 | positive (#) | 20/212<br>(9.4%) | 39/832<br>(4.7%) | 1.70 | 0.05 | positive (#) | 40/212<br>(18.9%) | 71/832<br>(8.5%) | 1.2 | 0.30 |
| polypharmacy | 0.99 | negative | 50/877<br>(5.7%) | 9/168<br>(5.4%) | 1.22 | 0.58 | positive (#) | 27/168<br>(5.7%) | 84/877<br>(5.4%) | 1.2 | 0.33 |
| eGFR | <0.001 | <42.0 (#) | 24/237<br>(10.1%) | 35/808<br>(4.3%) | 2.46 | <0.01 | ≥55.475 (#) | 91/474<br>(19.3%) | 20/571<br>(3.6%) | 3.9 | <0.001 |
| ALB | <0.001 | ≤4.3 (#) | 56/824<br>(6.3%) | 3/151<br>(2.0%) | 2.89 | 0.05 | ≤ 4.0 (#) | 50/343<br>(14.6%) | 61/647<br>(8.7%) | 2.0 | <0.001 |
| P1NP | 0.28 | ≤22.6 | 17/138<br>(12.3%) | 42/907<br>(4.6%) | 2.32 | <0.01 | ≤ 25.8 | 29/186<br>(15.6%) | 82/859<br>(9.5%) | 1.1 | 0.67 |
| TRACP-5b | 0.13 | ≤421 | 30/404<br>(7.4%) | 29/641<br>(4.5%) | 1.50 | 0.12 | ≤ 264 | 24/137<br>(17.5%) | 82/908<br>(9.0%) | 1.1 | 0.78 |
| Hgb | <0.001 | ≤12.0 (#) | 30/437<br>(6.9%) | 29/608<br>(4.8%) | 1.60 | 0.07 | ≤ 11.0 (#) | 50/343<br>(14.6%) | 61/702<br>(8.7%) | 2.0 | <0.001 |
| Lymph | <0.001 | ≥1,136 (#) | 50/751<br>(6.5%) | 9/270<br>(3.3%) | 1.80 | 0.10 | ≤ 1,423 (#) | 62/471<br>(13.2%) | 49/574<br>(8.5%) | 1.7 | <0.01 |
| Mono | 0.86 | >271 | 46/704<br>(6.5%) | 13/330<br>(3.8%) | 1.81 | 0.05 | > 218 | 104/890<br>(11.7%) | 7/155 (4.5%) | 1.9 | <0.05 |

### 2.1 Group comparison after PSM

Because the SQ grade showed a significantly higher risk ratio only for the incident VF, a PSM procedure was conducted for VFs to level the confounding factors. After the PSM, the dataset was narrowed to 560 patients, with 140 patients for each grade group. In these, 43 patients had incident VFs; 11 (7.9%), 9 (6.4%), 8 (5.7%), and 15 (10.7%) in the G-0, G-1, G-2 and G-3 were included, respectively. The confounding factors, such as CHF, C-I, MADS, and T-score in the LS, and eGFR_CysC, showed no statistical differences in any pairs of the SQ grade groups. However, VEC cannot diminish the statistical difference in any way because 100% of the patients in the G-3 group had the positivity of VEC. In the other factors, age at baseline, follow-up length, presence of AAC, DM, HL, administration of OPD, ALB, P1NP, Hgb, and Lymph showed significant differences between groups. The incidence of VF and NVF showed no significant difference between any pairs in the SQ grade (Table 7).

**Table 7:**
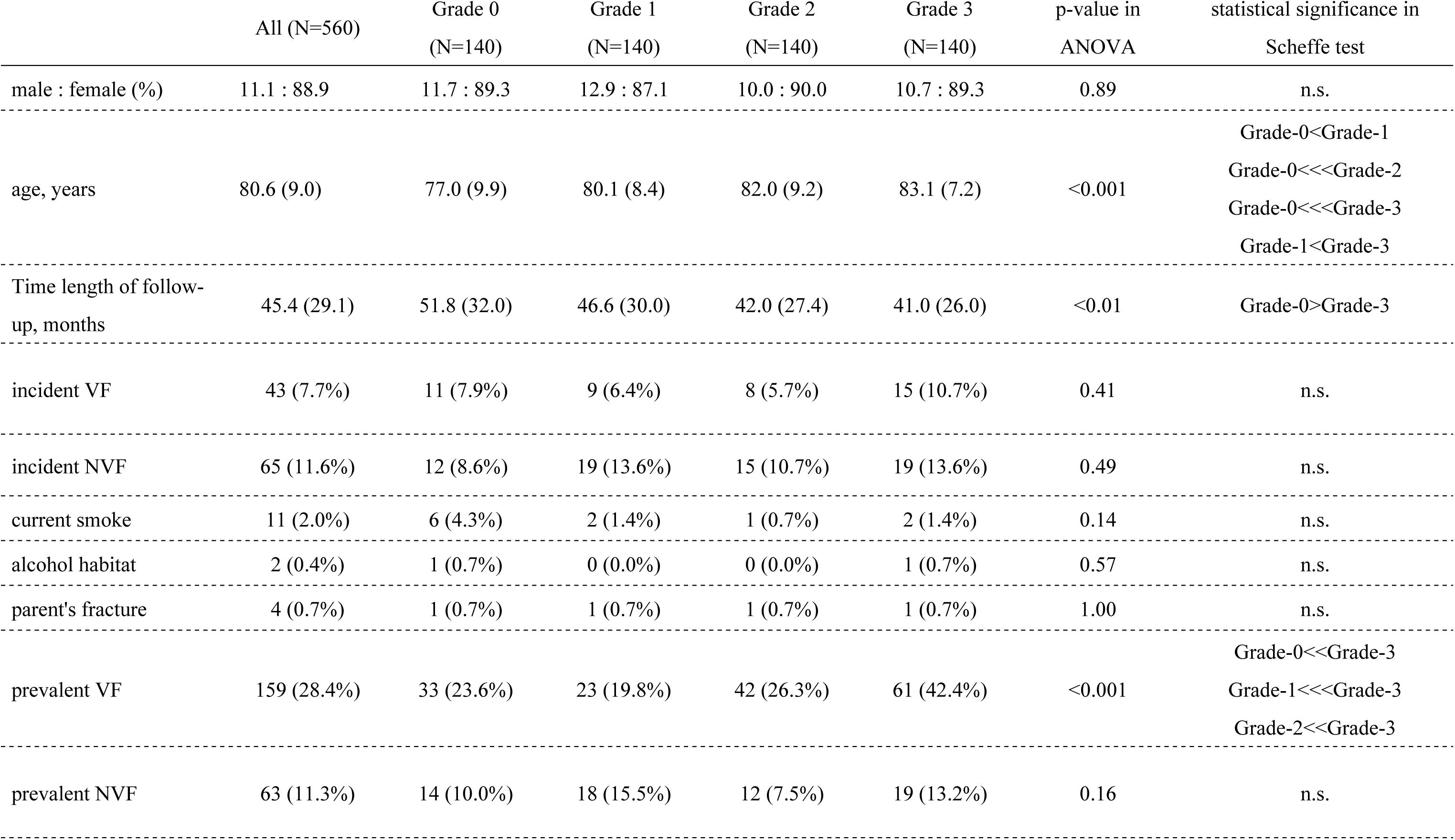
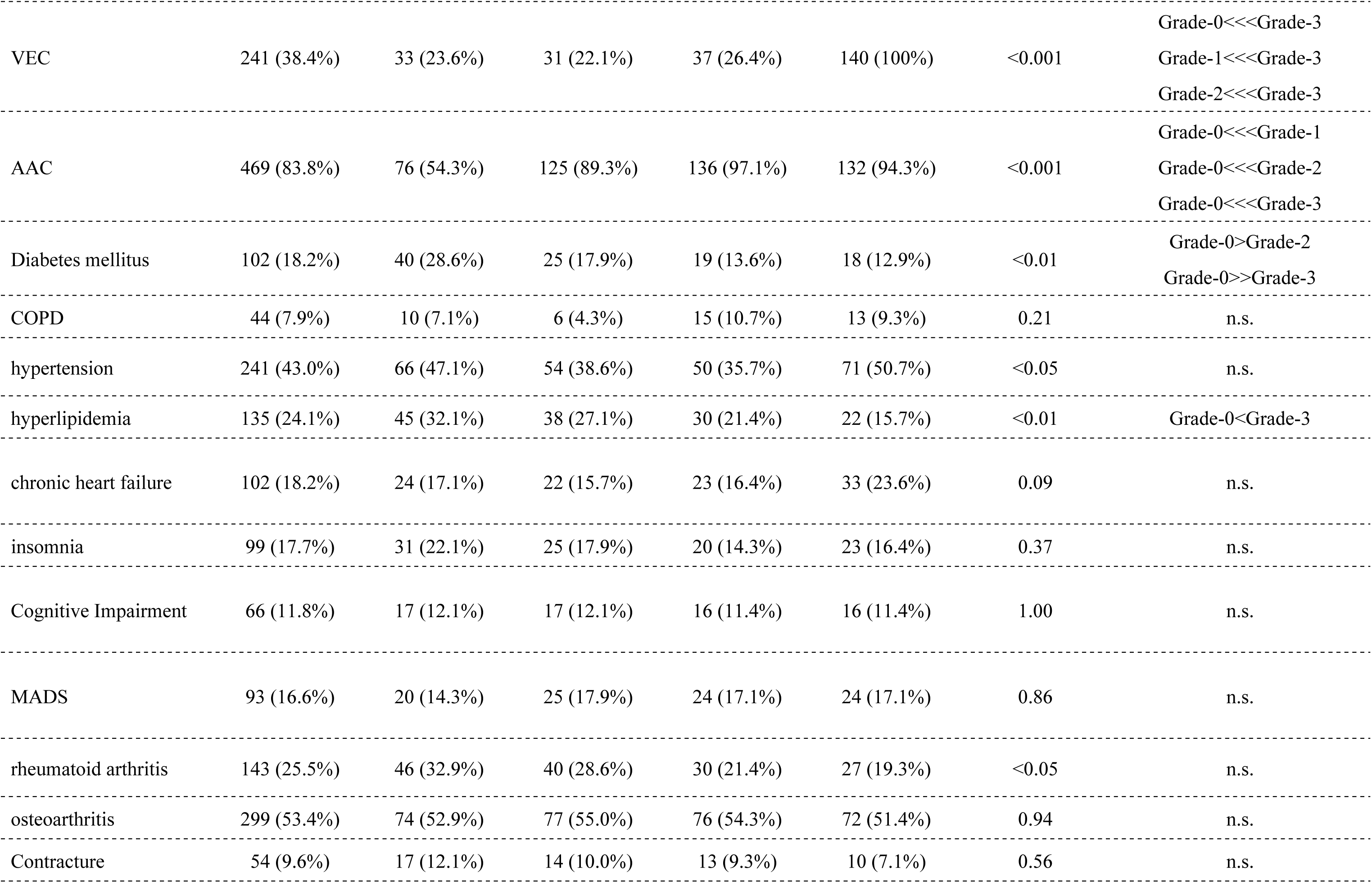
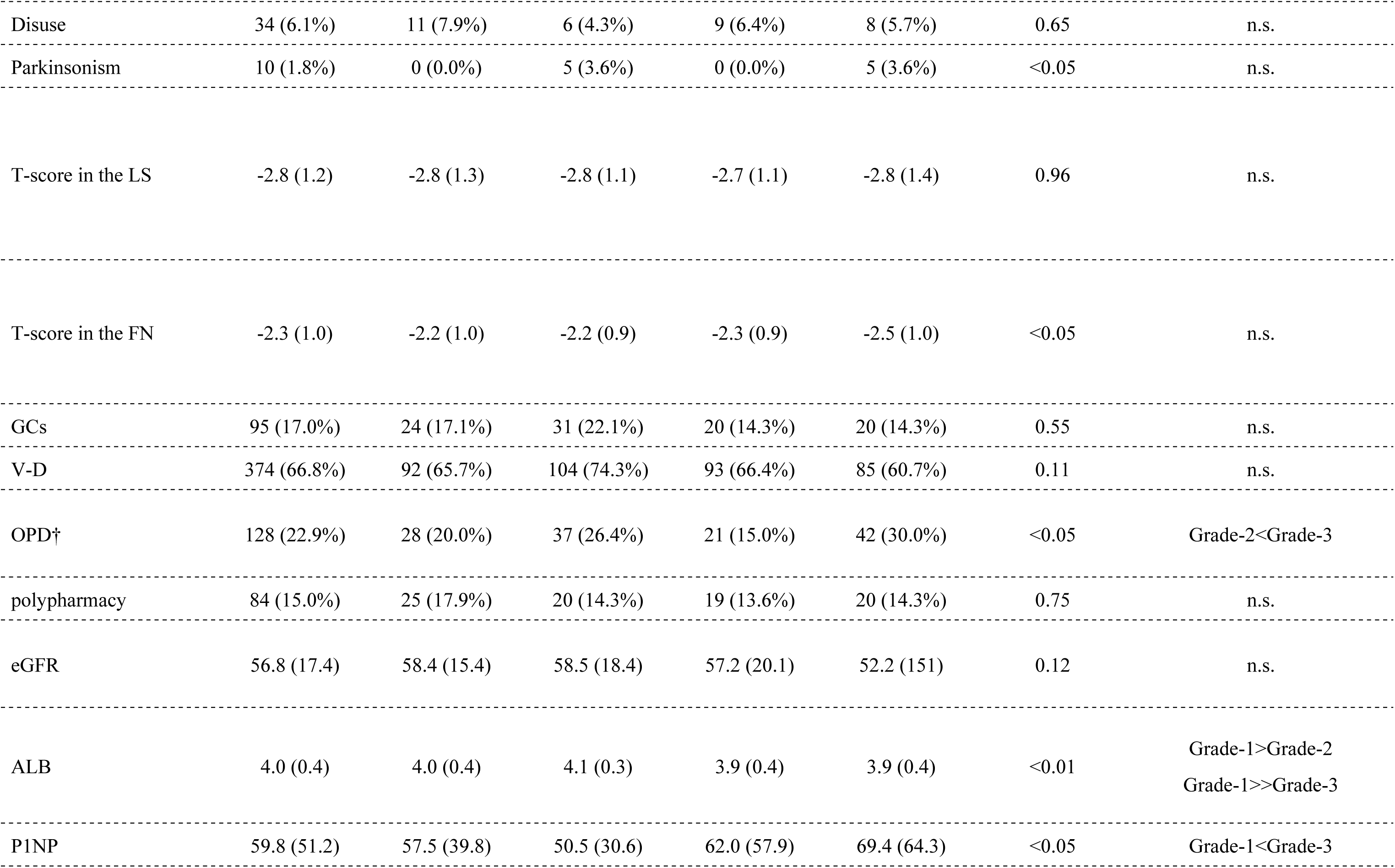
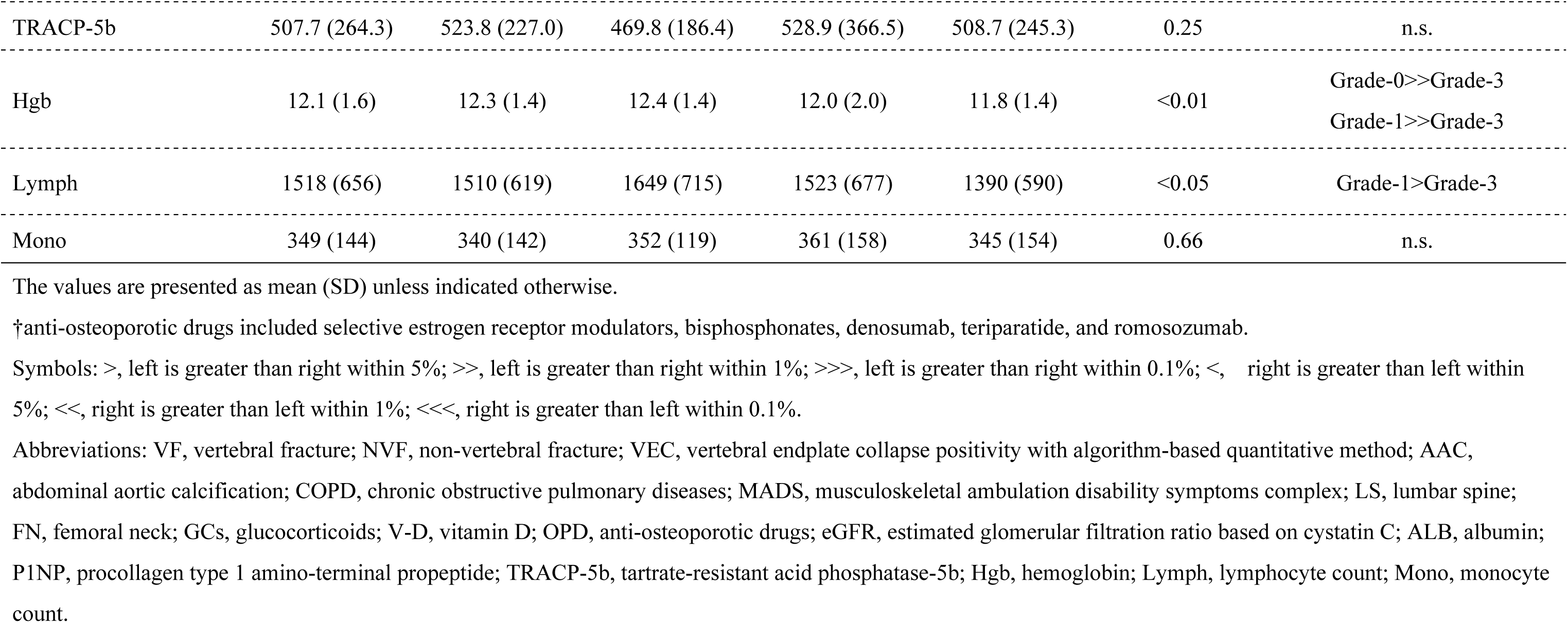
Demographic characteristics of the patient groups after the propensity score matching procedures.

In the Kaplan-Meier survival analysis, no pairs in the SQ grade showed significant Hazard ratios for an incident VF (Table 2). However, there was a significantly higher hazard ratio for an incident NVF in G-1 compared to G-0 and G-3 compared to G-0 (Table 3)

### 2.2 Risk factor analysis in the dataset after PSM

Older age and blood Lymph count were significant factors for the likelihood of the incident VFs using a Cox regression analysis with a univariate model in the dataset after PSM. However, the multivariate model showed no significant factors (Table 8).

**Table 8:**
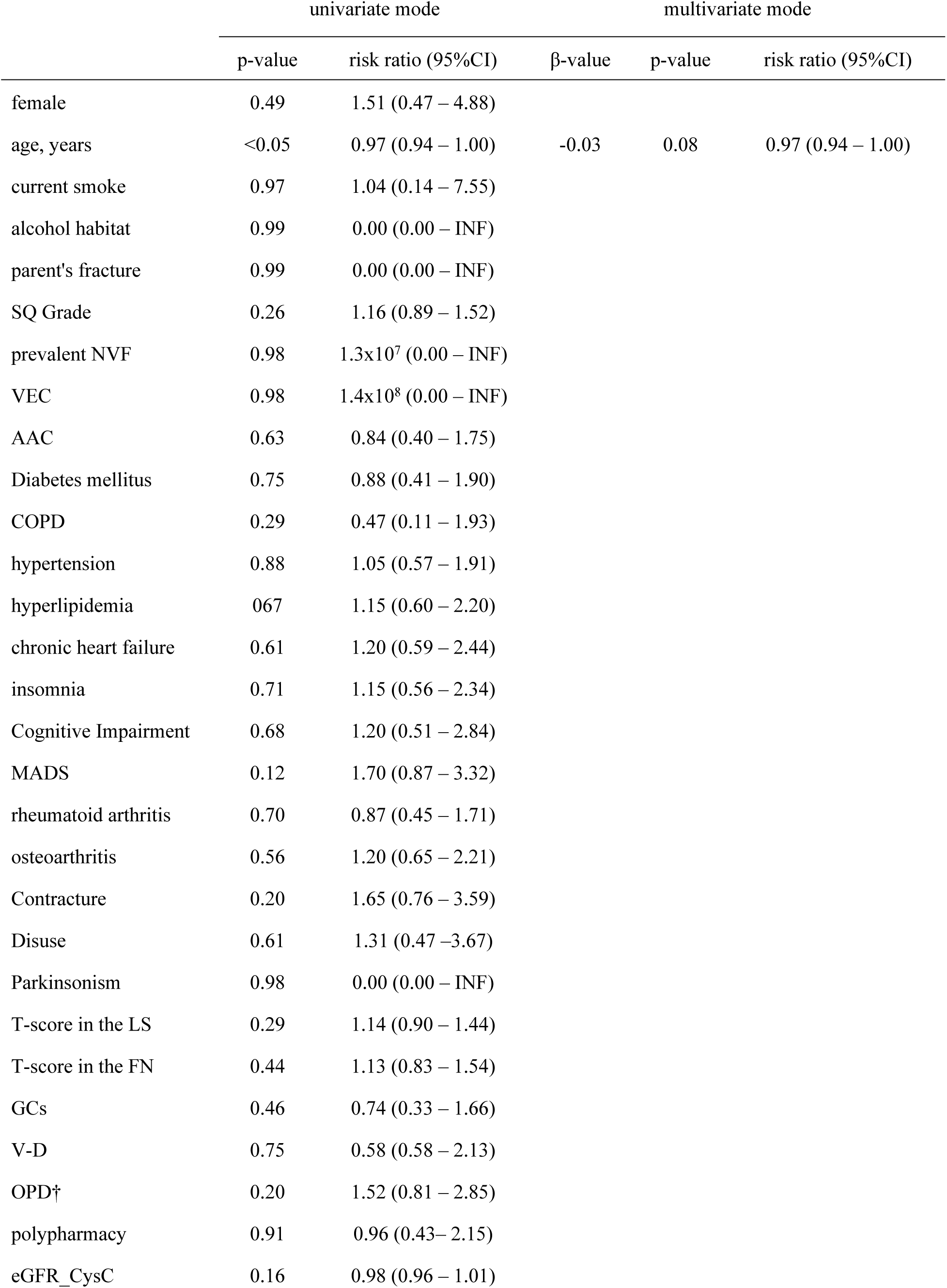
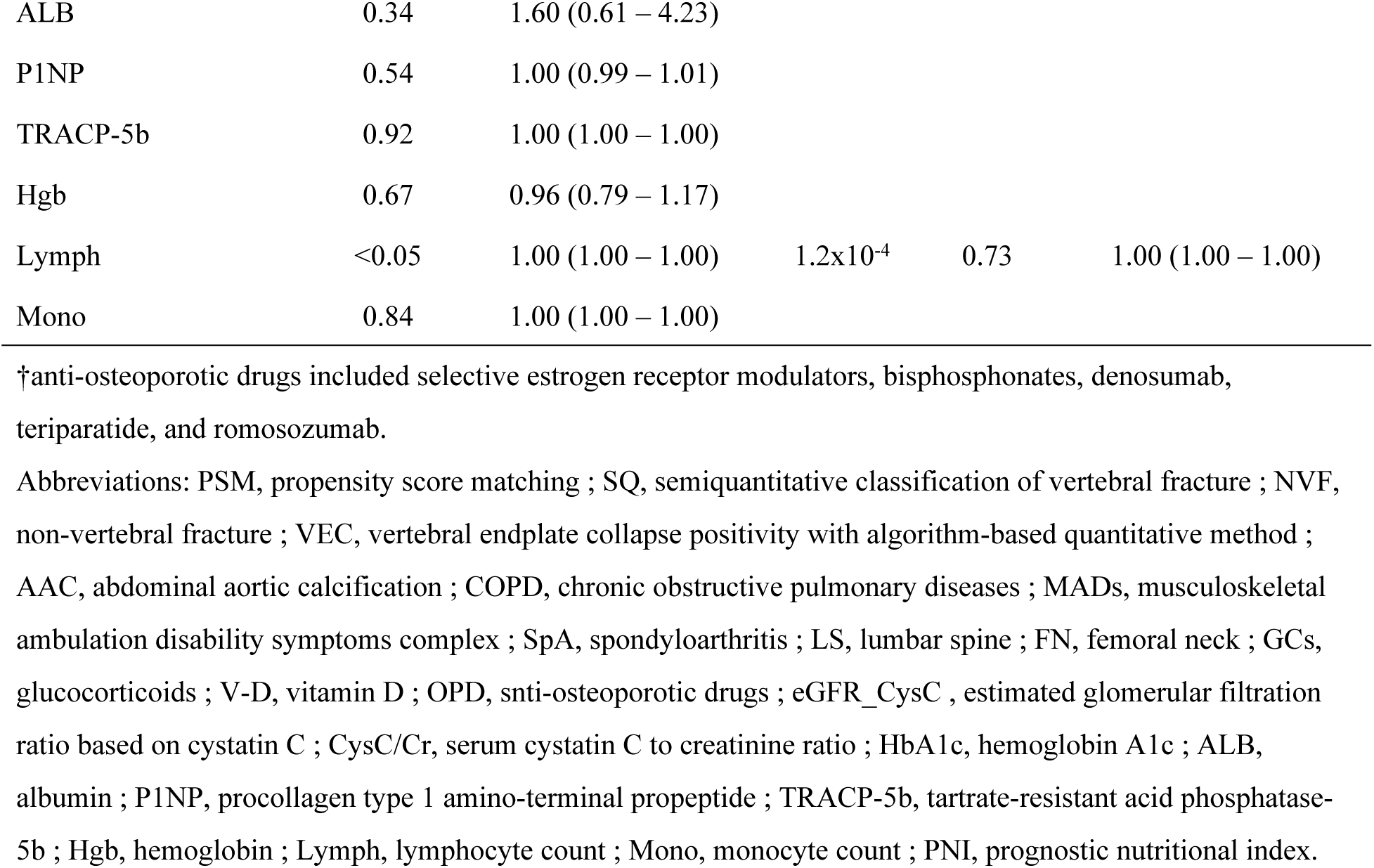
Results of a Cox regression analysis in the dataset after PSM.

| | univariate mode | | β-value | multivariate mode | |
| --- | --- | --- | --- | --- | --- |
|  | p-value | risk ratio (95%CI) |  | p-value | risk ratio (95%CI) |
| female | 0.49 | 1.51 (0.47 – 4.88) |  |  |  |
| age, years | <0.05 | 0.97 (0.94 – 1.00) | -0.03 | 0.08 | 0.97 (0.94 – 1.00) |
| current smoke | 0.97 | 1.04 (0.14 – 7.55) |  |  |  |
| alcohol habitat | 0.99 | 0.00 (0.00 – INF) |  |  |  |
| parent's fracture | 0.99 | 0.00 (0.00 – INF) |  |  |  |
| SQ Grade | 0.26 | 1.16 (0.89 – 1.52) |  |  |  |
| prevalent NVF | 0.98 | 1.3x10<sup>7</sup> (0.00 – INF) |  |  |  |
| VEC | 0.98 | 1.4x10<sup>8</sup> (0.00 – INF) |  |  |  |
| AAC | 0.63 | 0.84 (0.40 – 1.75) |  |  |  |
| Diabetes mellitus | 0.75 | 0.88 (0.41 – 1.90) |  |  |  |
| COPD | 0.29 | 0.47 (0.11 – 1.93) |  |  |  |
| hypertension | 0.88 | 1.05 (0.57 – 1.91) |  |  |  |
| hyperlipidemia | 0.67 | 1.15 (0.60 – 2.20) |  |  |  |
| chronic heart failure | 0.61 | 1.20 (0.59 – 2.44) |  |  |  |
| insomnia | 0.71 | 1.15 (0.56 – 2.34) |  |  |  |
| Cognitive Impairment | 0.68 | 1.20 (0.51 – 2.84) |  |  |  |
| MADS | 0.12 | 1.70 (0.87 – 3.32) |  |  |  |
| rheumatoid arthritis | 0.70 | 0.87 (0.45 – 1.71) |  |  |  |
| osteoarthritis | 0.56 | 1.20 (0.65 – 2.21) |  |  |  |
| Contracture | 0.20 | 1.65 (0.76 – 3.59) |  |  |  |
| Disuse | 0.61 | 1.31 (0.47 – 3.67) |  |  |  |
| Parkinsonism | 0.98 | 0.00 (0.00 – INF) |  |  |  |
| T-score in the LS | 0.29 | 1.14 (0.90 – 1.44) |  |  |  |
| T-score in the FN | 0.44 | 1.13 (0.83 – 1.54) |  |  |  |
| GCs | 0.46 | 0.74 (0.33 – 1.66) |  |  |  |
| V-D | 0.75 | 0.58 (0.58 – 2.13) |  |  |  |
| OPD† | 0.20 | 1.52 (0.81 – 2.85) |  |  |  |
| polypharmacy | 0.91 | 0.96 (0.43 – 2.15) |  |  |  |
| eGFR_CysC | 0.16 | 0.98 (0.96 – 1.01) |  |  |  |
| ALB | 0.34 | 1.60 (0.61 – 4.23) |  |  |  |
| P1NP | 0.54 | 1.00 (0.99 – 1.01) |  |  |  |
| TRACP-5b | 0.92 | 1.00 (1.00 – 1.00) |  |  |  |
| Hgb | 0.67 | 0.96 (0.79 – 1.17) |  |  |  |
| Lymph | <0.05 | 1.00 (1.00 – 1.00) | 1.2x10<sup>-4</sup> | 0.73 | 1.00 (1.00 – 1.00) |
| Mono | 0.84 | 1.00 (1.00 – 1.00) |  |  |  |

†anti-osteoporotic drugs included selective estrogen receptor modulators, bisphosphonates, denosumab, teriparatide, and romosozumab.
Abbreviations: PSM, propensity score matching ; SQ, semiquantitative classification of vertebral fracture ; NVF, non-vertebral fracture ; VEC, vertebral endplate collapse positivity with algorithm-based quantitative method ; AAC, abdominal aortic calcification ; COPD, chronic obstructive pulmonary diseases ; MADs, musculoskeletal ambulation disability symptoms complex ; SpA, spondyloarthritis ; LS, lumbar spine ; FN, femoral neck ; GCs, glucocorticoids ; V-D, vitamin D ; OPD, anti-osteoporotic drugs ; eGFR\_CysC , estimated glomerular filtration ratio based on cystatin C ; CysC/Cr, serum cystatin C to creatinine ratio ; HbA1c, hemoglobin A1c ; ALB, albumin ; P1NP, procollagen type 1 amino-terminal propeptide ; TRACP-5b, tartrate-resistant acid phosphatase-5b ; Hgb, hemoglobin ; Lymph, lymphocyte count ; Mono, monocyte count ; PNI, prognostic nutritional index.

### 3.1 Association between the significant factor and the SQ grade

The SQ grade was not a significant factor, but the presence of HT was the significant factor for incident NVF. Thus, binary logistic regression analysis was conducted between the presence of HT and the SQ grade. The SQ grade significantly correlated with the presence of HT (p<0.001). The probability of the incident NVF in the SQ grade group for the presence of HT was 10.0%, 19.4%, 21.7%, and 27.3% in Grade 0, Grade 1, Grade 2, and Grade 3, respectively, for the presence of the HT group. The probability of the incident VF was 3.1%, 3.9%, 7.5%, and 15.5%, respectively. On the other hand, in the HT absence group, the probabilities for the NVF were 2.3%, 3.4%, 4.9%, and 9.2%, and those for the VF were 3.1%, 2.3%, 4.9%, and 9.2% in Grade 0, Grade 1, Grade 2, and Grade 3, respectively. The NVF probability showed a stepwise increase according to the SQ grades, regardless of HT presence. When the patients were divided by an SQ grade of 0 or higher (SQ≥1), Cox regression analysis for an incident NVF examined SQ≥1 and HT, revealing that both factors showed significantly higher risk ratios of 3.73 and 3.10 for HT and SQ≥1 (in both cases, p<0.001), respectively.

## Discussion

The presence/absence of a prevalent fracture is one of the most decisive risk factors for subsequent osteoporotic fractures [16]. There are reports that the risk of subsequent VF increases by 4.4 times when there is an incident VF compared to when there is no VF [31], and this is the most crucial factor to pay attention to when considering osteoporosis treatment; therefore, prevalent fracture episode is adopted in the FRAX [32,33].

The SQ method is a screening method for VDs. It is widely known as an easy screening for VD. It is also widely used as a screening tool for predicting incident VFs and sometimes incident OFs, including NVF. It is considered an excellent method for screening because it allows judgment to be made without measuring bone mineral density. However, its classification is often the subject of debate. There is an undisputed consensus that SQ Grade 3, which is severe, is an apparent risk factor [34], but there are pros and cons as to whether SQ Grade 1 and Grade 2, which are mild and moderate, are risk factors. But does this consensus hold in the real-world clinical setting?

The number of incident VFs was less than the incident NVF, which is different from other established reports [34–37]. However, this study is conducted in a clinical-based population setting, whereas many previous reports used population-based studies. The mean age in our study is relatively high, which is one reason for a smaller number of VFs than NVFs, as the prevalence of hip fracture increases as age increases. Moreover, the number of cases with prevalent VF was recorded more than that with prevalent NVF.

So, it is a reasonable background data. Further, our study’s primary endpoint was only symptomatic clinical VFs, which should limit the number of cases with incident VFs. A global trend of fragility fractures also showed an overwhelming number of hip fractures relative to VFs [38].

In addition to SQ grade, we also considered previously reported potential items in extracting predicted risk factors [39]. We also considered bone metabolism markers. VEC positivity with ABQ is considered the most substantial vertebral body fracture factor. In the crude dataset used in the study, all 59 patients who presented incident VF had VEC positivity with ABQ. The results using Kruskal-Willis and Kaplan-Meier analyses show that the presence of VEC demonstrated statistically solid significance regarding the SQ grade and likelihood of the incident VFs and NVFs. The VEC using ABQ is a visual judgment of the discontinuity of bone contour, so the positive findings should mean prevalent vertebral body fracture; however, the findings cannot distinguish whether it is fresh or not, and this judgment is not the same as a clinical fracture, despite clinical fracture and the VEC are overlapped in considerable part. Therefore, the VEC positivity with ABQ is a strong indicator of the prediction of incident vertebral fracture, but it is not definitive.

The probability of incident VFs in Grade 3 was significantly higher than in the other three groups using the ANOVA Scheffe test; however, the statistical significance diminished with the Kaplan-Meier survival analysis. There was no significant difference in the likelihood of incident VFs in the Grade 1 and the Grade 2 groups compared to the Grade 0 group using the ANOVA Scheffe test. The results of the Kaplan-Meier survival analysis supported the significance of higher risk in the Grade 3 group. Yet, there was no statistical significance between the Grade 2 and Grade 3 groups; however, there was a statistically significant hazard ratio between the Grade 0 and Grade 2 using a Kaplan-Meier survival analysis. We examined a Cox regression analysis to extract significant factors without confounding elements. When a multivariate model was analyzed in Cox regression analysis, SQ Grade was the only factor with a significantly higher risk ratio. However, there are substantial differences in many background factors between groups, and there is a risk that these may act as confounding factors. Therefore, we needed to limit the groups using PSM procedures, leveling the confounding factors recognized and diminishing the influence of these factors.

On the other hand, using a Kaplan-Meier survival analysis, the probability of incident NVFs in Grade 1 or higher was significantly higher than in Grade 0. However, the SQ grade was not a significant risk factor; the presence of HT was the only significant factor using the multivariate model of Cox regression analysis. This could be explained by the fact that the SQ grade confounds HT.

After the PSM procedure, VEC could not be leveled because 100% of the patients in G-3 had positive VEC. However, the other factors were leveled. In the dataset after PSM, the SQ grade groups had no significant difference in the probability of incident clinical VFs and NVFs. Still, there were substantial differences in background factors, such as age, blood lymphocyte count, and PNI, despite these variables losing their statistical significance compared to the multivariate model. These could be ignored because the crude dataset showed no statistically significant higher risk ratios in these factors.

From the above results, it was found that SQ Grade classification has weak significance as a predictor of incident fracture, even including Grade 3. The SQ Grade, determined only by radiographs, needs help with accuracy. If higher precision is necessary, imaging methods such as high-resolution CT that can observe the destruction of the posterior wall on the vertebral body and damage to the endplate or other morphological index in the lumbar spine are desirable [23,40,41]. However, SQ grade is suitable for screening because it can be easily determined and is considered an excellent method for the initial evaluation before osteoporosis treatment.

Limitations include that it is a single-center study, that the follow-up period is short (average of less than four years), that it is a retrospective study, that the survey is statistically weak even though bias adjustment was performed. Another important thing is that this study is not a population-based analysis but a clinical-based analysis, so the sample size needed to be more significant for these many variables to analyze statistics. In the Cox regression analysis with a multivariate model in the crude dataset, 59 patients with incident VF and 111 with incident NVF were recorded. The number of variables that showed statistical significance was 11 for VF and 20 for NVF. These are too many for the sample size. And the underlying clinical background was too many lifestyle-related diseases and rheumatic diseases. We cannot deny the sample bias even though the risk develops higher, especially in Grade 3. This can serve as reference information when screening for osteoporosis.

## Conclusion

In conclusion, clinical research was performed to evaluate the validity of the SQ method as a risk factor for subsequent VF and NVF. Results showed that Grade 3 shows a significantly higher risk ratio for an incident VF. However, the risk ratio is modified by the confounding factors. After confounding factors were adjusted, the risk ratio was leveled. For incident NVF, the SQ grade is not a significant determinant factor, but the SQ was closely correlated with the presence of HT, which showed a significantly higher risk ratio. Grade 1 or higher shows a significantly higher risk ratio as HT is demonstrated. The SQ grade classification is an available method for screening for initial osteoporotic treatment.

## Data Availability

All relevant data are included in the manuscript and its supporting information files.

## Acknowledgment

The authors appreciated Ms. Kaoru Kuwabara, Sayori Masuoka, Eri Morichika, and Aoi Yoshida for their dedicated data collection. The authors also appreciate Ms. Saori Tamura’s identical X-ray picture techniques.

